# The Association between Endometriosis and Immunological diseases

**DOI:** 10.1101/2024.07.08.24310092

**Authors:** Nina Shigesi, Holly R. Harris, Hai Fang, Anne Ndungu, Matthew R. Lincoln, The International Endometriosis Genome Consortium, The 23andMe Research Team, Chris Cotsapas, Julian Knight, Stacey A. Missmer, Andrew P. Morris, Christian M. Becker, Nilufer Rahmioglu, Krina T. Zondervan

**Author notes:** Corresponding authors /. Jointly directed the work. Deceased (Anne Ndungu).

## Abstract

The evidence for a greater prevalence of immunological-diseases among endometriosis patients has varied in robustness and been subject to selection bias. We investigated the phenotypic and genetic association between endometriosis and 31 immunological-diseases in the UK Biobank (8,223 endometriosis, 64,620 immunological-disease cases). In cross-sectional and retrospective cohort analyses, endometriosis patients were at significantly increased (30-80%) risk of classical- autoimmune (rheumatoid arthritis, multiple sclerosis, coeliac disease), autoinflammatory (osteoarthritis) and mixed-pattern (psoriasis) diseases. Osteoarthritis (genetic-correlation (rg)=0.28, P=3.25×10^-15^), rheumatoid arthritis (rg=0.27, P=1.54×10^-5^) and multiple sclerosis (rg=0.09, P=4.00×10^-3^) were significantly genetically correlated with endometriosis. Mendelian randomisation analysis suggested a causal association between endometriosis and rheumatoid arthritis (OR=1.16, 95%CI=1.02-1.33). Expression QTL analyses highlighted effector genes enriched for seven pathways across all four conditions, with three genetic loci shared between endometriosis and osteoarthritis and one with rheumatoid arthritis. Although the increased risk of immunological-diseases among endometriosis patients is modest, their shared genetic basis opens-up opportunities for new treatments.

## Introduction

Endometriosis is a chronic inflammatory condition that features the presence of endometrial-like tissue in locations outside the uterus, mainly in the pelvic cavity^1^. The most commonly accepted explanation for the origin of the majority of these endometrial deposits is retrograde menstruation, when menstrual blood containing endometrial cells travels up the fallopian tubes into the pelvic cavity^2^. However, this phenomenon is experienced by the majority of menstruating individuals^3^, leaving the question why only in some endometrial cells are able to adhere to peritoneal surfaces, thrive, and proliferate^4^. Proliferation of the endometrial implants requires oestrogen, which is provided both systemically but also from localised production of aromatase^1^ and expression of oestrogen receptor beta^5^, inhibition of TNF-alpha induced apoptosis, increased interleukin-1beta which enhances cellular adhesion and proliferation, and localised inflammation^6^. Endometriotic implants secrete various cytokines, chemokines and prostaglandins, eliciting an inflammatory response that attracts macrophages, monocytes, neutrophils, T cells and eosinophils. Impairment of the innate, and possibly adaptive, immune system in removing ectopic endometrial cells from the peritoneal cavity appears to play a role in endometriosis^1^, ^7^. In particular, altered function of natural killer cells and macrophages has been implicated, but it is unclear if these aberrations play a role in causation or are part of pathophysiology^1^, ^8^.

Given the link with aberrant immune response, many clinical and population studies have investigated the association between endometriosis and auto-immune diseases, even postulating that endometriosis itself may be an auto-immune disorder^9–11^ because of the dysfunction in natural immunity. While auto-antibodies are not typically involved in the pathogenesis of endometriosis^12^ and thus it is not classified as an auto-immune condition^13^, a systematic review of published clinical and population studies suggested an increased risk of several autoimmune conditions (systemic lupus erythematosus, Sjögren’s syndrome, rheumatoid arthritis, autoimmune thyroid disorder, coeliac disease, multiple sclerosis, inflammatory bowel disease, and Addison’s disease) among females with endometriosis. However, most of the studies were limited by small sample sizes, selection biases, and lack of adjustment for confounding factors^14^.

Here we aim to investigate the association, and any shared biological basis, between endometriosis and 31 immunological disorders grouped into classical auto-immune, auto- inflammatory and mixed-pattern conditions^15^. We explore the association between endometriosis and immunological conditions in one of the largest available data sources, the UK Biobank data; conduct the largest possible genome-wide association study (GWAS) meta-analyses of conditions exhibiting significant phenotypic associations with endometriosis; use these datasets to investigate genetic correlations, potential causal pathways, and shared genetic risk variants between endometriosis and immunological diseases; and use endometrium and blood expression QTL data to identify genes dysregulated by shared disease-associated variants.

## Results

### Phenotypic association between endometriosis and immune conditions

The phenotypic association between endometriosis and immunological conditions was investigated in the UK Biobank (UKBB) using both cross-sectional and retrospective cohort study designs, with the latter assuming endometriosis as a diagnostic risk factor preceding an immunological disease diagnosis (see Methods). For the cross-sectional analysis, 8,223 endometriosis cases vs. 265,181 female controls without known endometriosis were included; and 64,620 immunological disease cases vs. 208,784 female controls without known immunological diseases. Supplementary Table 1 shows factors that were determined as potential confounders or mediators in the association analyses between endometriosis and immunological diseases. Adding factors significantly associated with both endometriosis and immunological diseases in a logistic regression model with endometriosis as exposure and any immunological disease as the outcome (see Methods), none were found to be confounders or mediators that significantly affected the effect size of association (>5% change). However, genetically determined ancestry and age at recruitment were included a-priori as potential confounders.

In both the cross-sectional and retrospective cohort analyses (Table 1), females with endometriosis vs. those without had a significantly increased risk for all immunological diseases combined (OR: 1.32 (1.25-1.39); HR: 1.32 (1.20-1.45)), classic autoimmune diseases (OR: 1.24 (1.13-1.36); HR: 1.41 (1.15-1.74)), autoinflammatory diseases (OR: 1.33 (1.26-1.41); HR: 1.29 (1.17-1.43)), and mixed-pattern diseases (OR: 1.23 (1.10-1.52); HR: 1.88 (1.25-2.81)).

**Table 1.**
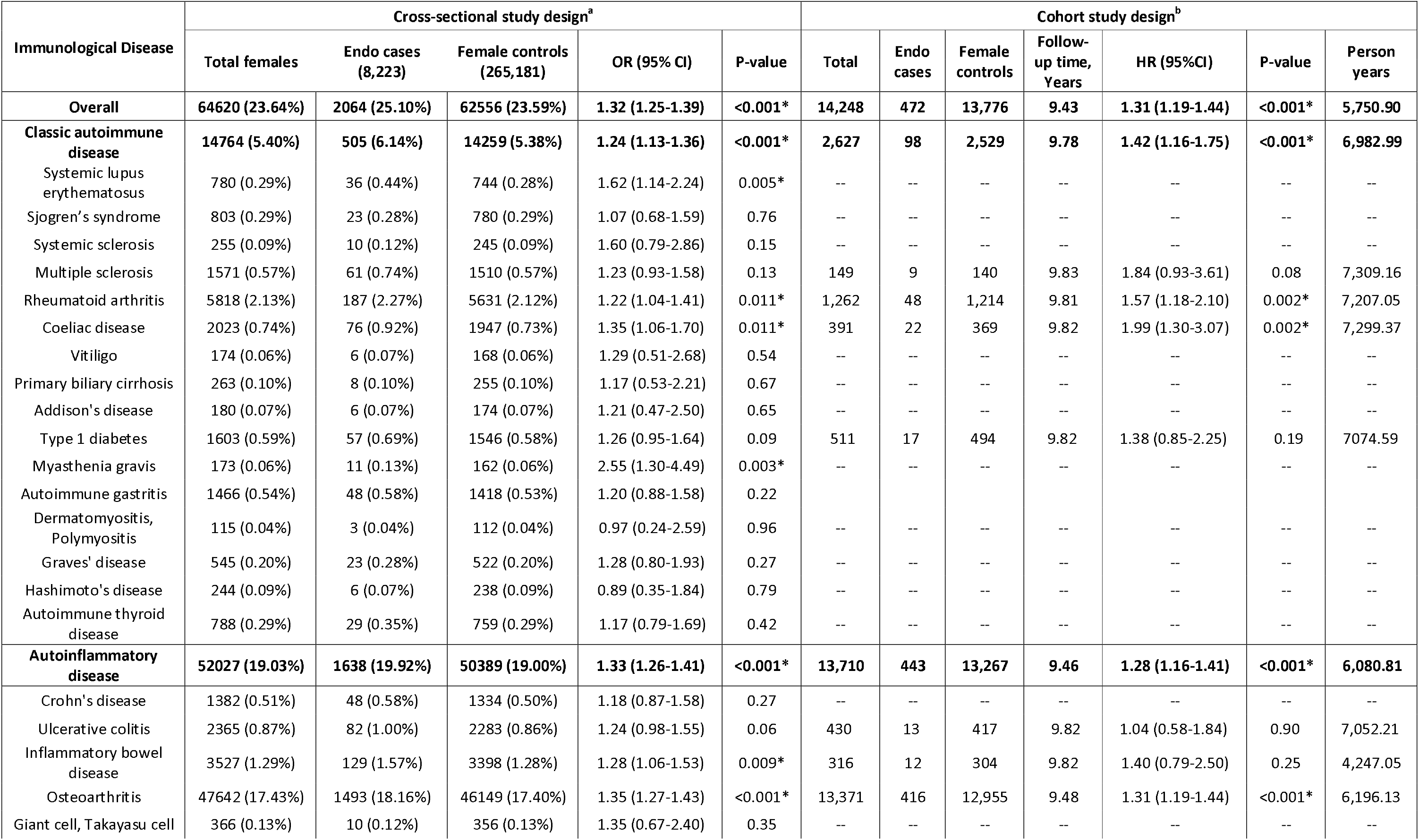
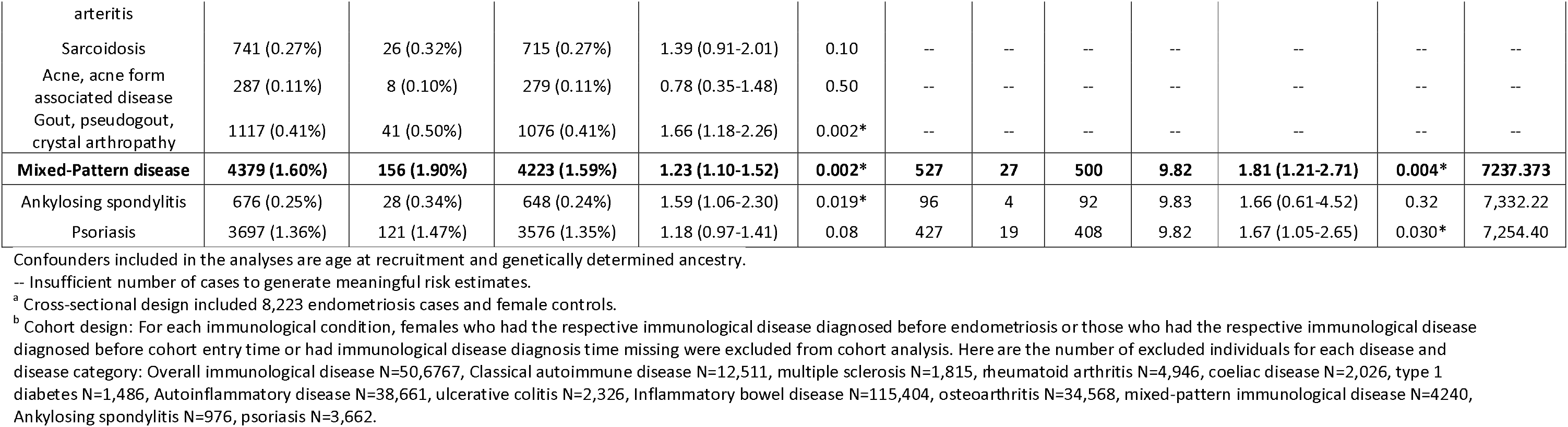
Immunological disease risks among females with vs. without endometriosis in UK Biobank utilising two different study designs: Cross-sectional study design (N=26 immunological disease with >100 female cases) and cohort study design (N=9 immunological disease with >1500 female cases)

Immunological diseases significantly associated with endometriosis in both cross-sectional and cohort analyses were: rheumatoid arthritis (OR:1.22 (1.04-1.41), P = 0.011; HR: 1.57 (1.18-2.10), P = 0.002); coeliac disease (OR: 1.35 (1.06-1.70), P = 0.011; HR: 1.99 (1.30-3.07), P = 0.002); and osteoarthritis (OR: 1.35 (1.27-1.43), P < 0.001; HR: 1.31 (1.19-1.44), P < 0.001). In addition, in the ‘gold standard’ cohort analyses, psoriasis (HR: 1.67 (1.05-2.65), P = 0.030) was significantly associated with endometriosis. One immunological condition significantly associated with endometriosis in cross-sectional analysis, systemic lupus erythematosus (OR:1.62 (1.14-2.24), P=0.005), myasthenia gravis (OR: 2.55 (1.30-4.49), P=0.003) and gout (OR:1.66 (1.18-2.26), P=0.002), could not be tested in a cohort study design due to insufficient case numbers. Overall, females with endometriosis compared to females without known endometriosis exhibited a 14% increased risk for at least having one immunological disease (OR = 1.14 (1.08-1.21)), a 21% increased risk for at least having two immunological diseases (OR = 1.21 (1.05-1.39)), and a 30% increased risk for having at least three immunological diseases (OR = 1.30 (0.92-1.78)) at any point in their lifetime (P < 0.001) (Supplementary Table 2).

When stratifying by menopausal status, gynaecological surgery (hysterectomy/oophorectomy), or hormone replacement therapy (HRT) use, effect sizes for the association between endometriosis and overall immunological disease risk remained largely unchanged (Supplementary Table 3).

### Genetic correlation between endometriosis and immunological diseases

For a total of eight immunological diseases associated with endometriosis either in cross-sectional or cohort analyses (ankylosing spondylitis, coeliac disease, inflammatory bowel disease, multiple sclerosis, osteoarthritis, psoriasis, rheumatoid arthritis, and systemic lupus erythematosus), we conducted female-only and sex-combined European ancestry GWAS analyses in UKBB (Table 2, See Methods). To achieve the greatest power to detect variants associated with each disease, sex-combined UKBB GWAS results were meta-analysed with existing GWAS summary statistics based on the largest sample sizes where available (Table 2), using the inverse variance weighted fixed-effects method as implemented in METAL (Supplementary Figures 1-8).

**Table 2.** Number of cases and controls of UKBB based female only and sex-combined GWAS for immunological conditions and summary of their meta-analysis with the largest publicly available European ancestry GWAS results.

| Immunological diseases | UKBB GWAS (Cases : Controls) |  | Published sex-combined GWAS (Cases : Controls) | Final sex-combined GWAS meta-analysis (Cases : Controls) |
| --- | --- | --- | --- | --- |
|  | Female-only | Sex-combined |  |  |
| Ankylosing spondylitis | 547 : 162,403 | 1,493 : 319,532 | N/A | 1,493 : 319,532 |
| Coeliac disease | 1,706 : 162,403 | 2,640 : 319,532 | 4,533 : 10,750<sup>72</sup> | 7,173 : 330,282 |
| Inflammatory bowel disease | 2,869 : 162,403 | 5,751 : 319,532 | 25,042 : 34,915<sup>54</sup> | 30,793 : 354,447 |
| Multiple sclerosis | 1,314 : 162,403 | 1,883 : 319,532 | 14,498 : 24,091<sup>56</sup> | 16,381 : 343,623 |
| Osteoarthritis | 39,866 : 162,403 | 68,878 : 319,532 | 77,052 : 378,169<sup>73</sup> | 77,052 : 378,169* |
| Psoriasis | 3,036 : 162,403 | 6,591 : 319,532 | 15,967 : 28,194<sup>59</sup> | 22,558 : 347,726 |
| Rheumatoid arthritis | 4,662 : 162,403 | 7,153 : 319,532 | 14,361 : 43,923<sup>57</sup> | 21,514 : 363,455 |
| Systemic Lupus erythematosus | 545 : 162,403 | 673 : 319,532 | 5,874 : 328,598<sup>74</sup> | 6,547 : 648,130 |

\* Published GWAS meta-analysis included UKBB hence, summary statistics from Tachmazidou et al.<sup>73</sup> were used in the analyses.

Utilising the GWAS meta-analysis summary results for the immunological diseases and the largest published endometriosis GWAS meta-analysis results (excluding UKBB GWAS results to achieve sample independence)^16^, we applied linkage disequilibrium (LD)-score regression (LDSC) analysis^17^, ^18^ to estimate the genetic correlation (rg) between endometriosis and the eight immunological diseases. Osteoarthritis (rg=0.29, p=3.25×10^-15^), rheumatoid arthritis (rg=0.26, p=1.54×10^-5^) and multiple sclerosis (rg=0.09, p=4.00×10^-3^) showed significant (p<6.25×10^-3^, see Methods) positive genetic correlations (rg) with endometriosis, suggesting a shared genetic component (Table 3).

**Table 3.** Genetic correlations from LDSC analysis between endometriosis and immunological disease. Multiple-testing correction for number of disease included in the analysis is applied (0.05/8=6.25×10^-3^) to determine significant correlations. UKBB endometriosis GWAS results were excluded from endometriosis meta-analysis to avoid overlap in LDSC analysis with immunological conditions for which we have analysed this dataset. H2: Heritability, SE: Standard error, RG: Genetic correlation.

| Immunological diseases | UKBB female-only GWAS |  |  | UKBB sex-combined GWAS |  |  | Sex-combined GWAS meta-analysis |  |  |
| --- | --- | --- | --- | --- | --- | --- | --- | --- | --- |
|  | H2 (SE) | RG (SE) | P-value | H2 (SE) | RG (SE) | P-value | H2 (SE) | RG (SE) | P-value |
| Ankylosing spondylitis | 0.006<br>(0.003) | 0.308<br>(0.124) | 0.013 | 0.003<br>(0.001) | 0.300<br>(0.124) | 0.016 | N/A | N/A | N/A |
| Coeliac disease | 0.013<br>(0.003) | 0.215<br>(0.083) | 0.010 | 0.009<br>(0.002) | 0.154<br>(0.068) | 0.024 | 0.077<br>(0.005) | 0.076<br>(0.054) | 0.1619 |
| Inflammatory bowel disease | 0.014<br>(0.003) | 0.054<br>(0.085) | 0.521 | 0.013<br>(0.002) | 0.007<br>(0.063) | 0.910 | 0.277<br>(0.028) | -0.041<br>(0.035) | 0.237 |
| Multiple sclerosis | 0.004<br>(0.003) | 0.247<br>(0.139) | 0.075 | 0.003<br>(0.001) | 0.273<br>(0.116) | 0.018 | 0.079<br>(0.005) | 0.088<br>(0.031) | $4.00 \times 10^{-3}$ |
| Osteoarthritis | 0.051<br>(0.004) | 0.322<br>(0.042) | $1.76 \times 10^{-14}$ | 0.042<br>(0.002) | 0.278<br>(0.038) | $2.46 \times 10^{-13}$ | 0.046<br>(0.002) | 0.278<br>(0.035) | $3.25 \times 10^{-15}$ |
| Psoriasis | 0.012<br>(0.003) | 0.043<br>(0.084) | 0.605 | 0.012<br>(0.002) | 0.049<br>(0.061) | 0.420 | 0.239<br>(0.027) | 0.066<br>(0.037) | 0.076 |
| Rheumatoid arthritis | 0.013<br>(0.003) | 0.277<br>(0.079) | 0.001 | 0.011<br>(0.002) | 0.284<br>(0.068) | $2.65 \times 10^{-5}$ | 0.064<br>(0.009) | 0.266<br>(0.062) | $1.54 \times 10^{-5}$ |
| Systemic lupus erythematosus | 0.003<br>(0.003) | 0.154<br>(0.154) | 0.316 | 0.002<br>(0.001) | 0.190<br>(0.165) | 0.248 | 0.332<br>(0.048) | 0.127<br>(0.071) | 0.074 |

### Mendelian randomisation (MR) analyses

MR analyses using genetic instrumental variables (IVs) were conducted to further investigate a potential causal relationship between endometriosis (exposure) and the increased risk of osteoarthritis, rheumatoid arthritis, and multiple sclerosis (outcomes) observed in the phenotypic association and genetic correlation analyses. Table 4 shows the results from the main MR-IVW model, with weighted median MR and MR-Egger regression provided as sensitivity analyses to test the robustness of the results. Analyses were conducted utilising 39 genome-wide significant (p<5×10^-8^) endometriosis associated LD-independent autosomal variants as IVs. Summary statistics for the IVs were extracted from UKBB female-only and meta-analysed sex-combined GWAS results to represent the outcomes of the three immunological diseases (see Methods).

**Table 4.** Mendelian Randomisation (MR) results for endometriosis vs. osteoarthritis, rheumatoid arthritis and multiple sclerosis.

| Immunological diseases | SNPs | Inverse variance weighted |  | Weighted median |  | MR Egger |  | MR Egger (P-values) |  |
| --- | --- | --- | --- | --- | --- | --- | --- | --- | --- |
|  |  | OR (95% CI) | P-value | OR (95% CI) | P-value | OR (95% CI) | P-value | Het<sup>a</sup> | Pleiotropy<sup>b</sup> |
| Osteoarthritis (39 IVs) |  |  |  |  |  |  |  |  |  |
| Female-only UKBB | 39 | 1.04 (0.97-1.11) | 0.282 | 1.05 (0.97-1.13) | 0.267 | 0.89 (0.72-1.07) | 0.203 | 0.002 | 0.084 |
| Female-only UKBB (outliers removed) | 35 | 1.03 (0.98-1.09) | 0.287 | 1.05 (0.97-1.13) | 0.250 | 0.95 (0.79-1.15) | 0.595 | 0.572 | 0.387 |
| Sex-combined meta-analysis | 39 | 1.02 (0.96-1.07) | 0.533 | 1.02 (0.97-1.08) | 0.400 | 1.08 (0.92-1.26) | 0.353 | 1.56x10<sup>-5</sup> | 0.229 |
| Sex-combined meta-analysis (outliers removed) | 36 | 1.01 (0.97-1.06) | 0.495 | 1.03 (0.98-1.09) | 0.281 | 1.03 (0.90-1.18) | 0.655 | 0.159 | 0.800 |
| Rheumatoid Arthritis (39 IVs) |  |  |  |  |  |  |  |  |  |
| Female-only UKBB | 39 | 1.16 (1.02-1.33) | 0.028 | 1.15 (0.94-1.40) | 0.169 | 1.15 (0.78-1.70) | 0.494 | 0.756 | 0.959 |
| Sex-combined meta-analysis | 31 | 1.06 (0.96-1.17) | 0.220 | 1.09 (0.94-1.26) | 0.261 | 1.08 (0.76-1.53) | 0.682 | 0.611 | 0.435 |
| Multiple Sclerosis (39 IVs) |  |  |  |  |  |  |  |  |  |
| Female-only UKBB | 39 | 1.12 (0.87-1.43) | 0.376 | 1.23 (0.87-1.73) | 0.243 | 1.75 (0.84-3.64) | 0.143 | 0.815 | 0.211 |
| Sex-combined meta-analysis | 39 | 1.06 (0.94-1.21) | 0.343 | 1.03 (0.89-1.20) | 0.692 | 1.24 (0.84-1.84) | 0.280 | 0.012* | 0.145 |
| Sex-combined Meta-analysis (outliers removed) | 38 | 1.04 (0.93-1.18) | 0.484 | 1.02 (0.88-1.2) | 0.759 | 1.20 (0.84-1.73) | 0.325 | 0.063 | 0.202 |

<sup>a</sup> Test for Heterogeneity, <sup>b</sup> Test for Pleiotropy, <sup>c</sup> Outlier IVs were identified by MR PRESSO software<sup>67</sup> (See methods and Supplementary Table 15).

For endometriosis vs. rheumatoid arthritis, utilising the 39 IVs illustrated a suggestive causal relationship between endometriosis vs. rheumatoid arthritis in females-only (OR [95%CI] = 1.16 [1.02-1.33], p-value=0.028). For endometriosis vs. osteoarthritis, and vs. multiple sclerosis, the MR analysis did not identify a significant causal relationship in either sex-combined or female-only analyses (Table 4).

### Multi-trait analysis of endometriosis and immunological diseases: osteoarthritis, rheumatoid arthritis and multiple sclerosis

To leverage the genetic sharing of association signals between endometriosis and osteoarthritis, rheumatoid arthritis, and multiple sclerosis for the discovery of additional endometriosis risk variants, we conducted a multi-trait analysis of GWAS (MTAG). MTAG capitalises on the genetic correlation between diseases to boost statistical power for detecting associations in genome-wide analyses^19^. The MTAG analysis was conducted for all four diseases simultaneously and identified 42 genome-wide significant lead SNPs for endometriosis (Supplementary Table 4), 6 of which were not reported previously^16^ (Supplementary Figure 9a-f). These 6 genetic variants are eQTLs for various genes across multiple tissues including *MSRA* and *PON2* protecting and repairing cells from oxidative stress in blood^20^, ^21^, *BLK* and *ZAP70* encoding enzymes belong to Tyrosine kinase family with roles in cell proliferation and differentiation in particular B-cell and T-cell development and adhesion^22^, ^23^, *ATRAID, SLC35F6, TMEM214* and *XKR6* involved in apoptosis-related pathways^24^, ^25^ and, *TRPS1* encoding a transcription factor that represses GATA-regulated genes involved in progesterone resistance and endometriosis progression in the pelvis^26^ (Supplementary Table 5).

The MTAG analysis for osteoarthritis yielded 27 significant variants (Supplementary Table 6), for rheumatoid arthritis yielded 28 significant variants (Supplementary Table 7) and for multiple sclerosis, it identified 64 genome-wide significant variants (Supplementary Table 8).

### Functional annotation of identified genome-wide significant variants and pathway analysis

We mapped genome-wide significant associations from each MTAG analysis for endometriosis, osteoarthritis, rheumatoid arthritis, and multiple sclerosis to the genes whose expression they are associated with, using GTEx V8 (54 tissues) and eQTLGen (31,684 blood datasets). We identified 439 genes regulated by 42 genome-wide significant endometriosis associated variants; 379 genes by 27 genome-wide significant osteoarthritis associated variants (Supplementary Table 6); 490 genes by 28 genome-wide significant rheumatoid arthritis variants (Supplementary Table 7); and 1,113 genes by 64 genome-wide significant multiple sclerosis associated variants (Supplementary Table 8). Of the 439 genes regulated by endometriosis risk variants, 192 were also regulated by a genome-wide significant risk variant of one or more of the other immune diseases (Figure 1).

**Figure 1.**
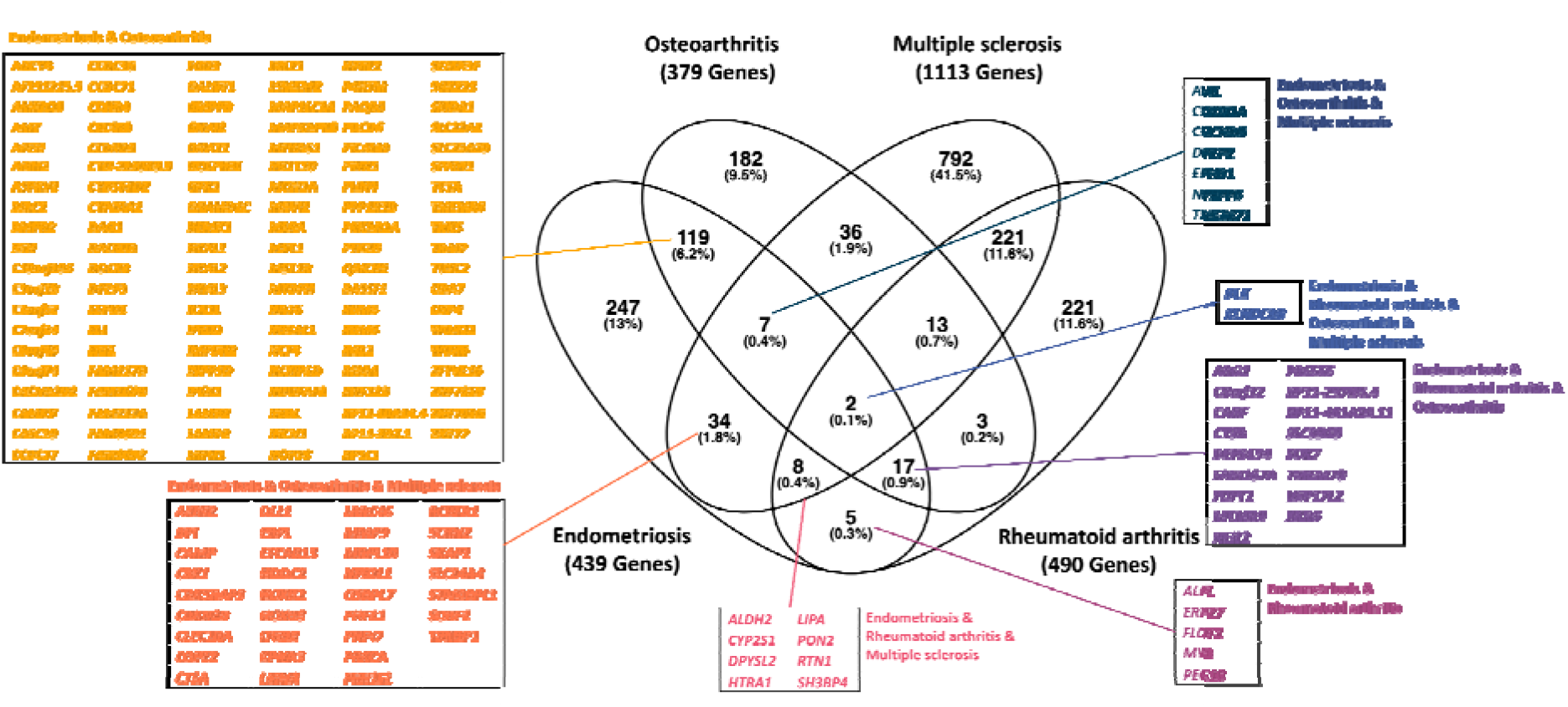
Overlap of genes associated with GWAS lead SNPs in eQTL analyses for endometriosis (42 genome-wide significant lead SNPs are eQTLs for 439 genes), osteoarthritis (27 genome-wide significant lead SNPs are eQTLs for 379 genes), multiple sclerosis (64 genome-wide significant lead SNPs are eQTLs for 1,113 genes) and rheumatoid arthritis (28 genome-wide significant lead SNPs are eQTLs for 490 genes) in various tissues in GTEx.

Pathway analysis (see Methods) based on the identified genes per disease identified numerous canonical pathways enriched with these genes (Supplementary Table 9-12). Among the top enriched pathways for endometriosis was ‘*signalling by receptor tyrosine kinases’,* a major class of cell surface proteins involved in signal transduction which triggers many downstream signalling pathways including *NFkB, MAPK* and *AKT*. These pathways are activated in endometriosis and have been suggested to harbour potential targets for non-hormonal therapeutics^27^.

Investigating the overlap of enriched genetically driven pathways between endometriosis, osteoarthritis, multiple sclerosis, and rheumatoid arthritis, we discovered that 45 out of the 79 enriched pathways for endometriosis were also enriched in the other immune conditions (Figure 2). In total 7 enriched pathways were shared across all four conditions, including ‘*signalling by receptor tyrosine kinases*’, ‘*innate immune system’*, ‘*adaptive immune system’*, ‘*extracellular matrix organisation’*, ‘*leukocyte trans-endothelial migration’*, ‘*lipid metabolism’*, and *‘arachidonic acid metabolism’* (Supplementary Figure 10). Within these overlapping enriched pathways, there are genes shared between conditions and also genes specific to each condition contributing to the pathway. For example, of the 21 genes enriched from endometriosis in ‘*signalling by reception tyrosine kinase*’, 8 are shared with osteoarthritis, including *NCF4, LAMB2, RHOA, MST1, MST1R, MAPKAPK3, DOCK3,* and *PTK2B,* and 3 are shared with multiple sclerosis including *ITGB3, PRKCA,* and *MMP9* (Supplementary Figure 10a). Another enriched pathway across the 4 conditions is ‘*arachidonic acid metabolism’*; of the 5 endometriosis genes enriched in this pathway, 4 are shared with the other 3 immune conditions, namely, *DPEP3, GPX1, DPEP2,* and *PON2 (Figure 2i)*.

**Figure 2.**
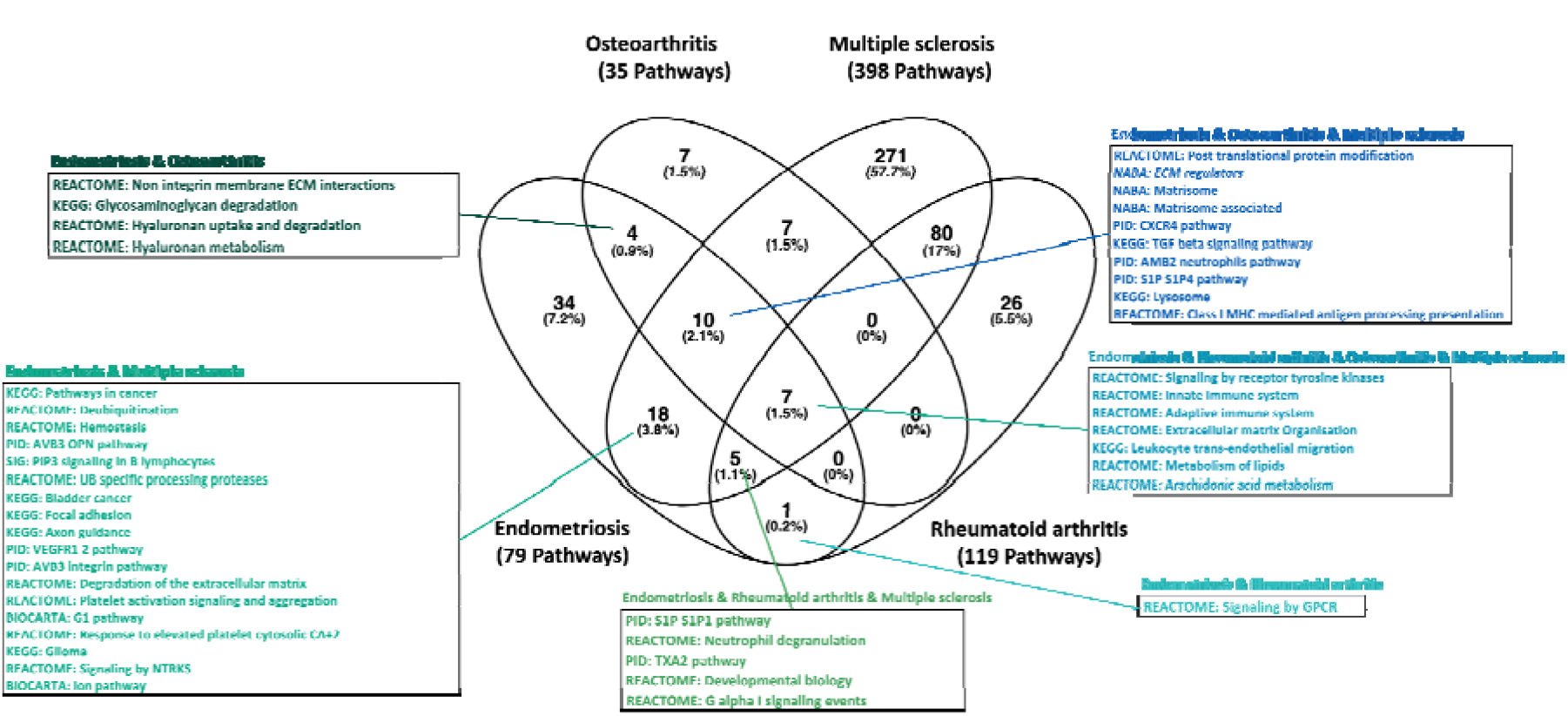
Overlap of pathways enriched with eQTL genes that are regulated by GWAS lead SNPs associated with endometriosis, osteoarthritis, rheumatoid arthritis and multiple sclerosis.

### Identification of shared genetic variants between endometriosis and immune diseases

A total of 12 osteoarthritis, rheumatoid arthritis and multiple sclerosis genome-wide significant lead SNPs were mapped within 1Mb of endometriosis genome-wide significant lead SNPs, with four of them tagging the same signal (r^2^>0.5) (Table 5, Supplementary Figure 9a-f). Three of these were shared with osteoarthritis (*BMPR2*/2q33.1, *BSN*/3p21.31, and *MLLT10*/10p12.31), and one with both osteoarthritis and rheumatoid arthritis (*XKR6*/8p23.1). MTAG association results of the 12 genome-wide significant endometriosis SNPs for osteoarthritis, rheumatoid arthritis and multiple sclerosis are provided in Supplementary Table 13.

**Table 5.**
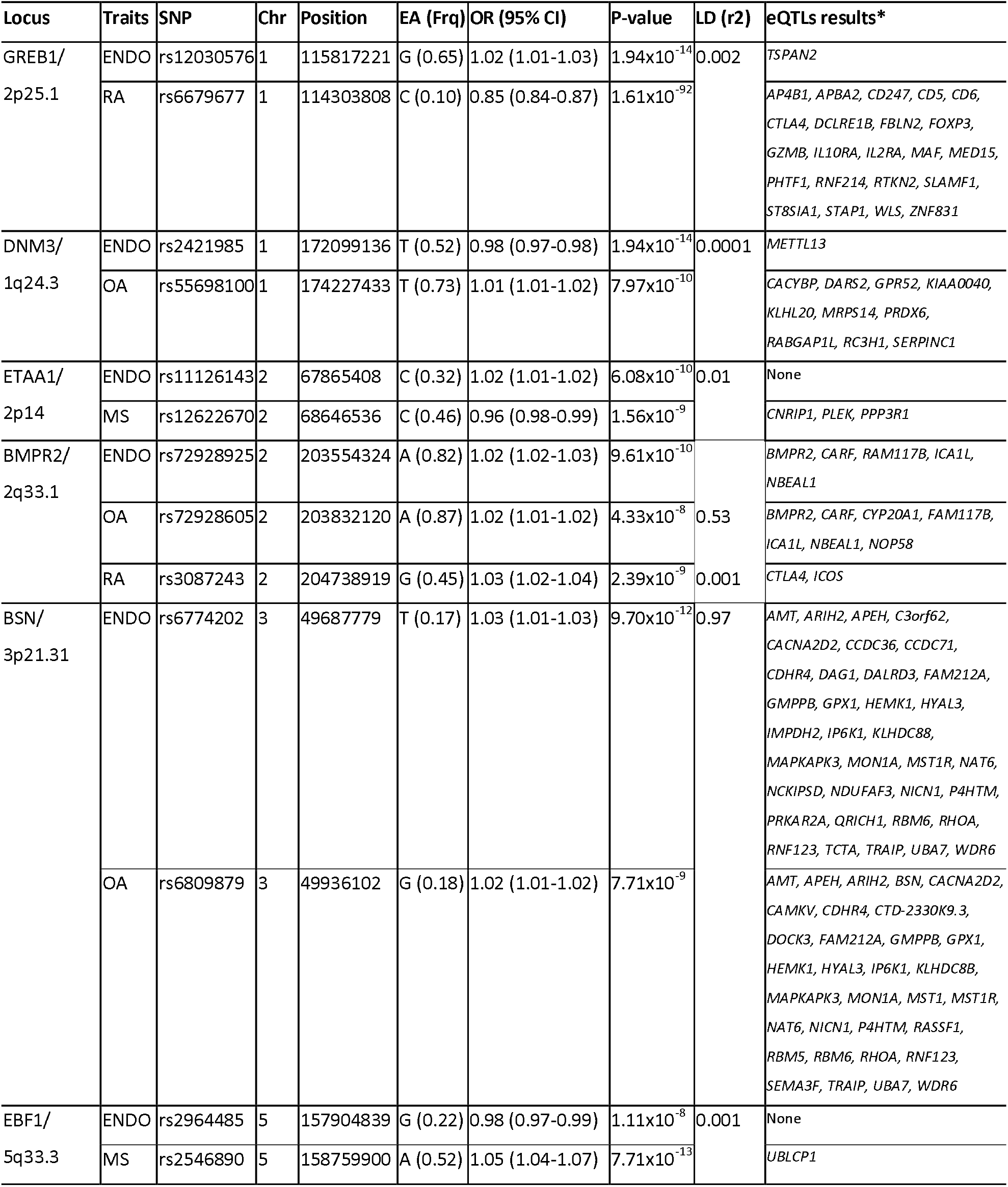
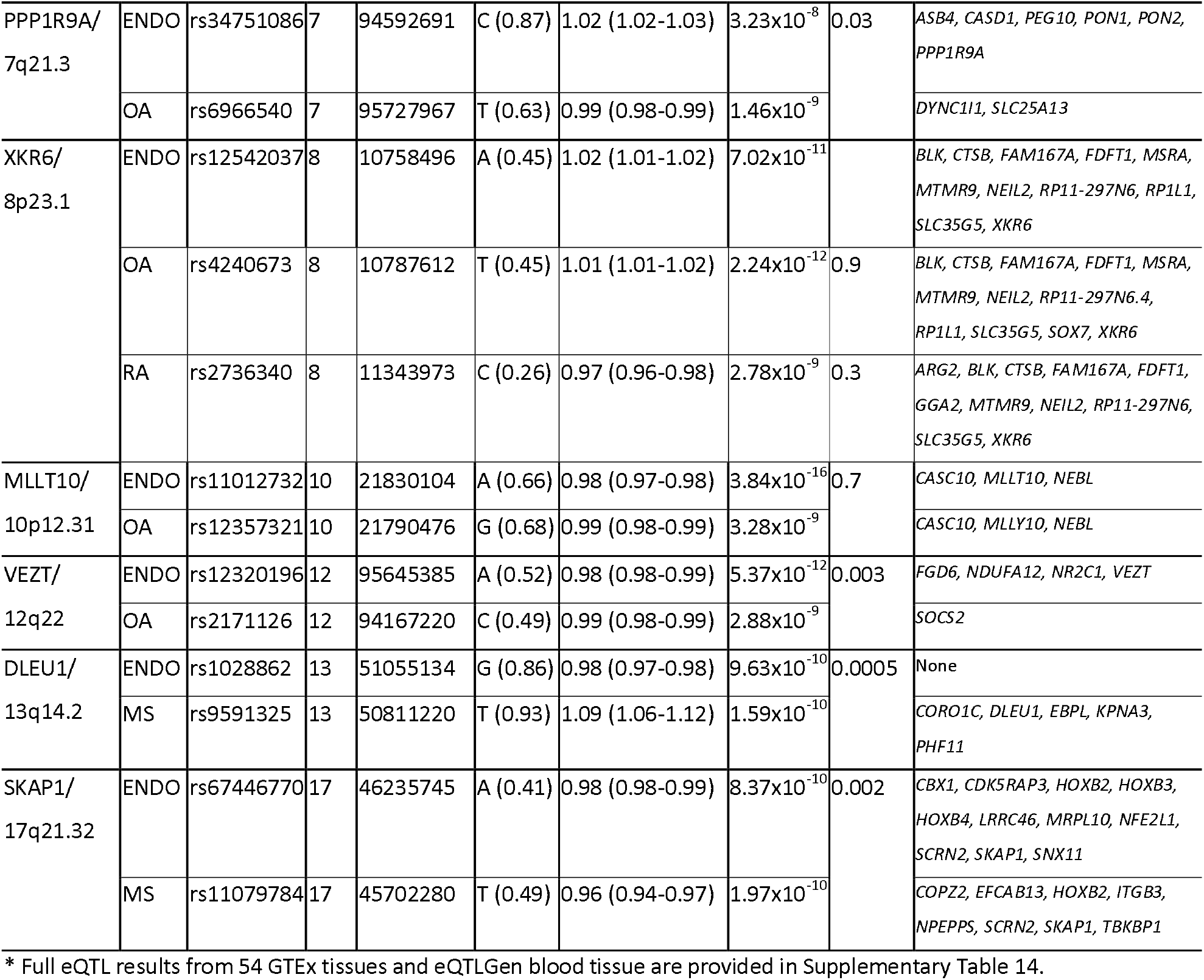
Genome-wide significant lead SNPs associated with endometriosis (ENDO) and rheumatoid arthritis (RA), osteoarthritis (OA) or multiple sclerosis (MS) that are located within 1Mb, with LD between them. Chr: Chromosome, AE: Effective allele, Frq: Effective allele frequency, LD: Linkage disequilibrium, eQTL: Expression quantitative trait loci.

At the *BMPR2/2q33.1* locus, the lead SNPs rs72928925 for endometriosis and rs72928605 for osteoarthritis are both eQTLs for *BMPR2* in blood and oesophagus muscularis (Supplementary Table 14). *BMPR2* encodes a member of the BMP receptor family of transmembrane serine/threonine kinases. The ligands of this receptor are members of the TGF-beta superfamily. The TGF-beta signalling pathway, involved in diverse cellular processes including cell proliferation, differentiation, apoptosis, and migration invasion, was also one of the pathways enriched with 10 eQTL genes regulated by endometriosis, osteoarthritis and multiple sclerosis associated variants (Figure 2, Supplementary Tables 9-12).

At the *BSN/3p21.31* locus, the lead SNP rs6774202 associated with endometriosis and rs6809879 with osteoarthritis are both eQTLs for a diverse set of overlapping genes (Table 5) that are part of pathways enriched between endometriosis and the other three immune conditions (Supplementary Tables 9-12). In particular, *RHOA* is part of the ‘*leukocyte trans-endothelial migration’* pathway that was enriched across all four conditions.

A third shared locus is *XKR6/8p23.1,* where the lead endometriosis SNP rs12542037 is in strong LD with the lead genome-wide significant osteoarthritis and rheumatoid arthritis SNPs (Table 5). This locus is involved in the regulation of multiple genes, namely *BLK*, *CTSB*, and *MTMP9,* which play roles in innate and adaptive immune system pathways^28^. In addition, *FDFT1,* regulated by the correlated genetic risk variants, encodes for squalene synthase that is involved in cholesterol biosynthesis. The ‘*lipid metabolism pathway’* is also enriched with genes regulated by genetic variants in each of the investigated 4 conditions (Supplementary Tables 9-12, Figure 2 and Supplementary Figure 10f).

The fourth locus previously implicated and described for endometriosis and osteoarthritis is *MLLT10*/10p12.31, which harbours genes such as *MLLT10* associated with pain perception and maintenance in multiple tissues^16^.

## Discussion

Our study of UKBB data reveals a significant increase in the risk of auto-immune and auto- inflammatory diseases among endometriosis patients, particularly in rheumatoid arthritis (HR:

1.57 (1.18-2.10), P = 0.002), coeliac disease (HR: 1.99 (1.30-3.07), P = 0.002), osteoarthritis (HR: 1.31 (1.19-1.44), P < 0.001), and psoriasis (HR: 1.67 (1.05-2.65), P = 0.030). Given the age at recruitment into UKBB (40-69 years, in 2006-10) and changes in awareness of endometriosis over time, the proportion of diagnosed females in UKBB (8,223 cases in 273,404 females = 0.03%) is relatively low compared to population prevalence estimates (up to 10%^1^) . This would have resulted in the undiagnosed females driving effect sizes for associations towards the null^29^, ^30^. Nevertheless, our results are consistent with evidence from previous case/control and cohort studies which showed significant association between endometriosis rheumatoid arthritis (RR:1.46 (0.70-3.03), coeliac disease (RR:1.39(1.14-1.70)), multiple sclerosis (OR:7.1 (4.4-11.3))^14^ and psoriasis (RR:1.75 (1.10-2.78))^31^. For systemic lupus erythematosus, cross-sectional evidence showed significant evidence (OR: 1.62 (1.14-2.24)) but we didn’t have enough cases to carry out the cohort study analysis. However, there is previous longitudinal evidence showing increased risk of systemic lupus erythematosus (HR:2.03 (1.17-3.51))^32^. Moreover, endometriosis patients compared to females without endometriosis were at increased risk of suffering from multiple immunological diseases which was most pronounced for autoinflammatory diseases (Supplementary table 2). This trend was observed in an early adulthood cohort study which we now expand to a broader age range.

Our genetic correlation analysis suggests that genetic factors contribute to the association between endometriosis and the increased risk of rheumatoid arthritis (rg=0.27, P=1.54×10^-5^), osteoarthritis (rg=0.28, P=3.25×10^-15^), and to a lesser extent, multiple sclerosis (rg=0.09, P=4.00×10^-3^). The significant genetic correlation between endometriosis and an immunological condition can be attributed to multiple mechanisms as investigated through MR analyses: (1) endometriosis causally leads to the subsequent development of the immunological condition; (2) endometriosis and the immunological condition share a common genetically driven cause; or (3) endometriosis and the immunological condition share multiple common causes, and the direction of effect between them can be complex^33^. Genetic correlation analyses are more powerful when the genetic architecture between conditions is polygenic involving many causal SNPs of small effect to leverage their aggregated effects, which is the case for endometriosis and the immunological conditions we studied here.

The MR analyses yielded no robust evidence of causal relationships between endometriosis and genetically correlated immunological conditions, except for a suggestive causal effect of endometriosis on rheumatoid arthritis in females-only (OR=1.16, 95% CI=1.02-1.33, p=0.028). MR analysis provides insights into whether the association between two complex conditions is causal by utilising genetic variants associated with the exposure (endometriosis). However, it is assumed that the genetic variants utilised as IVs have strong predictive power of the exposure, and the recommendation is to limit these to genome-wide significant (GWS) associated variants. However, even GWS variants as instruments often modestly predict the exposure, which can limit the power of MR analysis ^34^. The 39 endometriosis associated variants included as IVs in our MR analyses explain <2% of heritable variation (5% of stage III/IV disease^16^) which has implications for the interpretation of non-significant MR results. Our MR instruments would have been weighted towards risk for stage III/IV disease, in particular ovarian endometrioma^16^. Previous studies associating risk of auto-immune and inflammatory conditions with endometriosis included predominantly stage I/II cases^32^, ^35^, although some of this evidence was based on adolescents who may have been genetically predisposed to develop stage III/IV disease later in life^35^. Another limitation we encounter is a lack of available female-specific immunological disease GWAS meta-analysis results, which is surprising given that many immunological conditions exhibit higher prevalence in females. We conducted female-specific GWAS analyses in the UK Biobank, however, sample sizes were limited compared to sex-combined GWAS meta-analysis for these conditions in the literature.

The suggestive causal effect of endometriosis on rheumatoid arthritis is intriguing and warrants further exploration in the future, with IVs that explain a greater proportion of the genetic variability for endometriosis. This will require larger endometriosis GWAS to uncover more genetic variants contributing to the polygenic component of endometriosis. A large-scale female-specific rheumatoid arthritis GWAS meta-analysis should also result in a more relevant dataset to explore its potential causal basis with endometriosis. While power limitations hamper the interpretation of causal relationships between endometriosis and osteoarthritis, rheumatoid arthritis, or multiple sclerosis, results of the genetic correlation analyses highlight a shared genetic basis.

Understanding the basis of genetic sharing regardless of causality is important, as shared biological mechanisms of pathogenesis and pathophysiology could open new avenues for treatment development.

The MTAG analyses, leveraging genetic correlations between endometriosis, osteoarthritis, rheumatoid arthritis and multiple sclerosis, identified 42 genome-wide significant loci for endometriosis, 6 of which were not identified before: *ABHD1/*2p23.3, *TMEM131/*2q11.2, *XRCC4/*5q14.2, *PPP1R9A/*7q21.3, *XKR6/*8p23.1 and *TRPS1/*8p23.3. These variants were eQTLs for various genes involved in protecting and repairing cells from oxidative stress, in B-cell and T-cell development, apoptosis-related pathways and regulation of progesterone resistance. The MTAG-derived 42 GWS lead endometriosis-associated variants were mapped to 439 genes, 27 GWS lead osteoarthritis SNPs to 379 genes, 28 GWS lead rheumatoid arthritis SNPs to 490 genes and 64 GWS lead multiple sclerosis variants to 1113 genes. When we considered the overlap between these genes, 43.7% of endometriosis eQTL genes were shared with at least one of the three immunological conditions; the majority with osteoarthritis (33%) followed by multiple sclerosis (11.6%) and rheumatoid arthritis (7%). Pathway analysis revealed that 50.6% of pathways enriched in gene lists for endometriosis were also enriched in multiple sclerosis, 26.6% in osteoarthritis and 16.5% in rheumatoid arthritis. Seven pathways were enriched across endometriosis and all three immune conditions, including large, general immune regulatory pathways ‘*innate immune system’* (1100 genes) and ‘*adaptive immune system’* (807 genes). A more specific shared pathway was ‘*signalling by receptor tyrosine kinases*’. Receptor tyrosine kinases are a large family of cell-surface receptors that involved wide-variety inter and intra-cellular signalling. Previous studies have suggested the involvement of kinase signalling pathways and potential non-hormonal treatment targets therein for endometriosis^27^, which our analyses support. Another shared pathway, ‘*extracellular matrix organisation’*, included *MMP9, PRKCA and ITGB3. MMP9* encodes for a metalloproteinase that has a purported role in the progression of invasion in endometriosis as well as angiogenesis and fibrosis^36^, has involvement in a variety of inflammatory autoimmune diseases, and has been suggested to be a therapeutic target for autoimmune conditions^37^, ^38^.

*PRKCA* is involved in immune cell trafficking and *ITGB3* is coding for integrin beta3 expression of which is associated with autoimmune conditions including multiple sclerosis^39^.

Intriguingly,‘*arachidonic acid metabolism’* was another shared pathway, including four genes shared between endometriosis and osteoarthritis, rheumatoid arthritis and multiple sclerosis (*DPEP3, GPX1, DPEP2* and *PON2)*. Arachidonic acid is an essential fatty acid ingested through diet. Arachidonic acid derived prostaglandins contribute to inflammation through their role as intercellular pro-inflammatory mediators, and promote excitability of the peripheral somatosensory system contributing to pain exacerbation^40^.

Among the GWAS loci and effectors genes we identified through MTAG and eQTL analyses, three were shared between endometriosis and osteoarthritis (*BMPR2*/2q33.1, *BSN*/3p21.31, and *MLLT10*/10p12.31), and one, *XKR6*/8p23.1, between endometriosis and rheumatoid arthritis. *MLLT10* has been associated with pain perception and maintenance across multiple tissues and has been previously described^16^. *BMPR2* encodes a member of BMP receptor family of transmembrane serine/threonine kinases, acting as receptor for the TGF-beta superfamily. SNPs at the *BSN/3p21.31* locus are eQTLs for various genes including *RHOA*, part of the ‘*leukocyte trans- endothelial migration’* pathway that is enriched across all four conditions. Leukocytes migrate from blood into tissues as part of inflammation and immune surveillance. During this process leukocytes bind to cell adhesion molecules and migrate across the vascular endothelium. Another interesting eQTL gene associated with variants at *BSN/3p21.31* is *HYAL3,* which is involved in the hyaluronan/hyaluronic acid metabolism and glycosaminoglycan degradation pathways. Hyaluronic acid is a naturally occurring glycosaminoglycan most abundant in the extracellular matrix involved in various physiological processes including wound healing, tissue regeneration and joint lubrication. It is also used for relief of joint pain, wound healing and various other applications, and has been shown to reduce production of proinflammatory mediators, reduce sensitivity associated with osteoarthritis pain^41^, ^42^. Recent in-vivo and in-vitro studies suggest that hyaluronic acid may have the ability to reduce endometriosis lesion size in mice but that it may also promote inflammation when administered acutely^43^, suggesting further research into mechanism of action and therapeutic potential in endometriosis is needed.

*XKR6/8p23.1,* shared between endometriosis and rheumatoid arthritis, was associated with the regulation of multiple genes including *BLK*, *CTSB*, and *MTMP9* - which play roles in innate and adaptive immune system pathways^28^ – and *FDFT1,* involved in cholesterol biosynthesis.

Cholesterol is a precursor of steroid hormones and essential part of plasma membranes. It is also enriched in lipid rafts which play an important part in many cellular processes including signal transduction pathways, membrane trafficking, cytoskeletal organisation, apoptosis, cell adhesions and migration^44^. In the context of inflammatory conditions, lipid metabolism has been suggested to harbour targets for reducing inflammation without the undesirable side-effects of anti-inflammatory therapies^45^.

As mentioned, our analyses were limited by the lack of large-scale female-specific GWAS meta-analyses for immune conditions, particularly those exhibiting higher prevalence in females. It is well established that sex-specific genetic signatures are present for conditions showing variability by sex^46^, and female-specific GWAS results for immunological conditions may offer increased genetic correlations with endometriosis and opportunities for discovery and shared genetic signals. Future genetic comorbidity analyses should also explore results for different endometriosis subtypes. Recent GWAS analyses have suggested that ovarian endometriosis has a different genetic basis to peritoneal disease^16^, but the sample sizes for which summary statistics were generated did not allow for sufficiently powered inclusion in the present analyses. Similarly, future analyses should explore signals for different subtypes of immunological diseases, such as osteoarthritis^47^, once larger GWAS datasets become available. Lastly, genetic analyses were limited to European ancestry individuals and larger GWAS across more diverse ancestry groups are needed.

In conclusion, our results show that females with endometriosis are at a modestly (30-40%) increased risk of both auto-immune and auto-inflammatory conditions, and that comorbidity with osteoarthritis, rheumatoid arthritis, and to a more limited extent multiple sclerosis, is biologically underpinned. In terms of clinical relevance, we suggest awareness among treating physicians of this increased risk of comorbidity, in order to spot early symptoms of immunological conditions among females with endometriosis, and vice versa. While current clinical action is limited to increased vigilance, the results offer a wide range of novel avenues and targets for exploring mechanisms and potential cross-condition treatment development or repurposing.

## Methods

### Phenotypic analysis study population and disease ascertainment

The UK Biobank (UKBB) is comprised of 500K individuals aged 40-69 at time of recruitment (2006- 2010) from across the UK. The biobank was approved by the North West Multi-Centre Research Ethics Committee (MREC). In the UKBB, information was collected from participants during recruitment using questionnaires on socioeconomic status, behavioural, family history and medical history. Participants were also followed up for cause-specific morbidity and mortality through linkage to disease registries, death registries, hospital admission records and primary care data. In addition, a range of biological samples including blood, urine and saliva were collected from the participants. A more detailed description of the UKBB can be found in the UK Biobank protocol^48^.

Given that endometriosis is a gynaecological condition affecting those assigned female at birth, only the individual’s assigned female at birth (N=273,404) were included in the phenotypic association analysis with the immunological conditions. From this point onwards we will refer to those assigned female at birth as females in this manuscript. Endometriosis was identified based on self-reported data from questionnaires and/or hospital records (ICD10/9: N801-809 and 617.1- 9). A total 31 immunological conditions were identified from self-reported data and/or hospital records (ICD10/9) that were classified into three groups (94) as following: (1) autoinflammatory conditions: Acne (ICD10/9: L70* and 7060, 7061), acute respiratory distress syndrome (ICD10/9: J80, P220, 769, 769.9), erythema nodosum (ICD10/9: L52, 6952), giant cell/Takayasu arteritis (ICD10/9: M314, M315, M316, 4465, 4467), gout/pseudogout (ICD10/9: M10*, M11*, 274, 2740, 2741, 2748, 2749, 712, 7120, 7121, 7122, 7123, 7128, 71280, 71281, 71282, 71283, 71284, 71285, 71286, 71287, 71288, 71289, 7129, 71290, 71291, 71292, 71293, 71294, 71295, 71296, 71297, 71298, 71299), total inflammatory bowel disease (ICD10/9: K50, K500, K501, K508, K509, K51, K510, K511, K512, K513, K514, K515, K518, K519, 555, 5550, 5551, 5552, 5559, 556, 5560, 5569), Crohn’s disease (ICD10/9: K50, K500, K501, K508, K509, 555, 5550, 5551, 5552, 5559) Ulcerative colitis (ICD10/9: K51, K510, K511, K512, K513, K514, K515, K518, K519, 556, 5560, 5569), osteoarthritis (ICD10/9: M15, M150, M1500, M151, M152, M153, M154, M158, M159, M1599, M16, M160, M161, M162, M163, M164, M165, M166, M167, M169, M17, M170, M171, M172, M173, M174, M175, M179, M18, M180, M181, M182, M183, M184, M185, M189, M19, M190, M1900, M1911, M1912, M1913, M1914, M1915, M1916, M1917, M1918, M1919, M192, M1920, M1921, M1922, M1923, M1924, M1925, M1926, M1927, M1928, M1929, M198, M1980, M1981, M1982, M1983, M1984, M1985, M1986, M1987, M1988, M1989, M199, M19990, M1991, M1992, M1993, M1994, M1995, M1996, M1997, M1998, M1999, 715, 7150, 7151, 71510, 71511, 71512, 71513, 71514, 71515, 71516, 71517, 71518, 71519, 7152, 71520, 71521, 71522, 71523, 71524, 71525, 71526, 71527, 71528, 71529, 7153, 71530, 71531, 71532, 71533, 71534, 71535, 71536, 71537, 71538, 71539, 7158, 7159), sarcoidosis (ICD10/9: D860, D861, D862, D863, D868, D869, 135, 1359), (2) classical autoimmune conditions: Addison’s disease (ICD10/9: E271, E272, 25540, 25542), autoimmune gastritis(ICD10/9: D510, 2810), autoimmune thyroid disease (ICD10/9: E050, E063, 2420, 2452), Graves’ disease (ICD10/9: E050, 2420), Hashimoto’s disease (ICD10/9: E063, 2452), coeliac disease (ICD10/9: K900, 5790), dermatomyositis/polymyositis (ICD10/9: M33*, M360, 7103, 7104), multiple sclerosis (ICD10/9: G35, 340), myasthenia gravis (ICD10/9: G70, G700, G701, G702, G708, G709, 358, 3580, 35800, 35801, 35809, 3581, 3582, 3588, 3589), pemphigus/pemphigoid (ICD10/9: H133, L10, L100, L101, L102, L103, L104, L105, L108, L109, L12, L120, L121, L122, L123, L128, L129, 6944, 6945, 6946), primary biliary cirrhosis (ICD10/9: K743, 5716), rheumatoid arthritis (ICD10/9: M05*, M06*, 714, 7140, 71400-71409, 7141, 71410-71419, 7142, 71420-71429), Sjögren’s syndrome (ICD10/9: M350, 7102), systemic lupus erythematosus (ICD10/9: M32*, L90*, 6954, 7100), systemic sclerosis (ICD10/9: M34*, 5172, 7101), type 1 diabetes (ICD10/9: E10*, 25001, 25011, 25021, 25091), vitiligo (ICD10/9: L80, 70901), (3) Combination of autoinflammatory and autoimmune condition categories: Ankylosing spondylitis (ICD10/9: M081*, M45*, 7200), Behcet’s syndrome (ICD10/9: M352, 1361, 7112), reactive arthritis (ICD10/9: M023*, 0993, 7111), psoriasis/psoriatic arthritis/psoriatic arthropathies (ICD10/9: L40*, M070*, M073*, 6961, 6960). A common control set was defined as females without endometriosis diagnosis excluding anyone with diagnoses of any of the 31 immunological conditions.

Potential confounding or mediating factors were determined including age of recruitment, genetically determined ancestry, menopause status, age at menarche, parity, body size, BMI and fat distribution^49^, alcohol consumption, smoking, infertility and disease such as ovarian cancer^50^ and cardiovascular disease^51^, which have been illustrated to be associated with endometriosis and some immunological conditions. Age at recruitment (which represents potential age-related cohort effects) and ancestry were considered a-priori variables to be included in the models. Many of the other factors were assessed only at baseline recruitment into UKBB, which for most females would have followed rather than coincided with, or preceded, an endometriosis diagnosis, and therefore the potential for confounding vs. mediation effects could not be accurately assessed.

Nevertheless, to assess their potential impact on the associations, factors associated both with endometriosis and immunological disease were include in a logistic regression model with endometriosis as exposure and immunological disease as outcome. None of these factors either showed >5% change in effect (potential confounders) or removal of effects (mediators), and therefore only a-priori variables age at recruitment and genetically determined ancestry were included in the models.

### Phenotypic association analysis

Phenotypic association analysis between endometriosis and immune conditions was conducted utilising two different analysis methods: (1) a cross-sectional analysis to test for a simple association between risk of an immunological disease diagnosis with a diagnosis of endometriosis at any point in time, including all females in the UKBB; (2) a ‘gold standard’ cohort study design to incorporate temporality between diagnoses, where entry time was defined as the recruitment date into UKBB. Cross-sectional analysis was conducted for 26 immunological disease that had at least 100 female cases in UK Biobank. A total of 5 immune conditions were excluded from analysis due to number of cases <100: Reactive arthritis (N=4), Behcet’s syndrome (N=27), acute respiratory distress syndrome (N=79), erythema nodosum associated disease (N=79), pemphigus/pemphigoid (N=95). Cohort analysis was conducted for 9 immunological diseases with a minimum of 1,500 female cases to allow enough number of immunological disease cases after excluding prevalent immunological diseases diagnosed before cohort entry time or before the endometriosis diagnosis. The majority of females had immunological diseases diagnosed after endometriosis (66.8%, 1,275 out of 1,909 females with both diagnoses, had an immunological disease diagnosis after their endometriosis diagnosis). Therefore, endometriosis was treated as the exposure and immunological disease as the outcome in the cohort analyses. This also fits with the observation that many individuals with endometriosis have symptom onset in their teens or twenties, often many years before their ultimate diagnosis^52^. Females with an endometriosis diagnosis at the time of recruitment were classified as exposed, whereas those who had not had an endometriosis diagnosis at the time of recruitment were classified as unexposed. Those individuals who received an endometriosis diagnosis during follow-up, prior to any immunological disease diagnosis, contributed person-time to the unexposed group until the occurrence of endometriosis diagnosis, if any, and subsequently to the exposed group after diagnosis. For each immunological disease, females who had the respective immunological disease diagnosed before endometriosis or those who had the respective immunological disease diagnosed before cohort entry time or had immunological disease diagnosis time missing were excluded from cohort analysis (Table 1).

In the cross-sectional study analysis, the prevalence of each specific and categorized immunological diseases in females with and without a history of endometriosis diagnosis was investigated using logistic regression models with odds ratios (ORs) as risk measure. Cross-sectional study analysis for each specific and categorized immunological diseases was conducted with adjustment of age and genetically determined ancestry. In the cohort study, the risk of incident immunological diseases in females with and without endometriosis history was investigated using Cox proportional hazards regression models with calculated hazard ratios (HRs). The proportional hazards assumption was tested by function of “cox.zph” in the “survival” R library. In the cohort analysis, time to event was formulated from entry to the cohort until the end of follow-up time. The follow-up time (rather than age) is used as the underlying time variable, since the date of assessment is described in more detail with information on the exact date and months participants attended the assessment centre (to be used as the index date) in the UK Biobank. The end of follow-up time is the date of incident immunological diseases, death, loss to follow-up or end of follow-up (end date of follow-up is the date of last download of the dataset, which is 8^th^ Jan. 2019), whichever occurred first. Cohort analysis for each specific and categorized immunological diseases was conducted with adjustment of age (categorical: <50, 50-60, >=60) and genetically determined ancestry (categorical: white, non-white). All risk estimates were reported with 95% confidence intervals (CIs) and two-sided P-values. Person-years and mean follow-up time for each cohort analysis were calculated. All analyses were carried out using R software.

### Genome-wide association study (GWAS) and meta-analysis for immune conditions

Only genetically determined European ancestry individuals were included in the analysis. GWAS was conducted using UKBB data for females-only and sex-combined study population for 8 immune conditions: inflammatory bowel disease (N = 2,869 females; N = 5,751 sex-combined), osteoarthritis (N = 39,866 females; N = 68,878 sex-combined), ankylosing spondylitis (N = 547 females, N = 1,493 sex-combined), psoriasis (N = 3,036 females, N = 6,591 sex-combined), coeliac disease (N = 1,706 females, N = 2,640 sex-combined), multiple sclerosis (N = 1,314 females, N = 1,883 sex-combined), rheumatoid arthritis (N = 4,662 females, N = 7,153 sex-combined) and systemic lupus erythematosus (N = 545 females, N = 673 sex-combined). Controls were defined as a common control set without any diagnosis of immunological diseases or endometriosis within UKBB. The linear mixed model (LMM) implemented in BOLT^53^ was utilised for GWAS analysis to take into account relatedness in the data and to increase power of analysis by a linear mixed effects model. GWAS results were adjusted for a binary variable denoting the genotyping chip (the UKBB Axiom array or the UK BiLEVE array), for SNPs with minimum MAF filter of < 1% and included SNPs with ≤ 60% missingness.

Furthermore, published largest European ancestry GWAS results on these 8 immunological conditions were identified through literature^47^, ^54^–^60^ and downloaded for meta-analysis with UKBB GWAS results. Before meta-analysis, GWAS study-level QC was performed and markers absent in the 1000G reference panel, large missing values (≥ 60%) or lack beta/odds ratio estimates were excluded. GWAS meta-analysis for each immunological disease was carried out using METAL software using an inverse variance weighted fixed effect meta-analysis method^61^. GWAS meta-analysis results were filtered and excluded those with MAF < 1%, high heterogeneity (HetISquare > 90), present < 50% effective sample size (Neff; Neff = 4NCases*NControls/(NCases + NControls).

All genetic analyses were done on genome reference of homo sapiens (human) genome assembly GRCh37 (hg19). Genetic information on chromosome X was excluded. MHC region of Chr6:24000000-35000000 was excluded as it has a dense clustering of immune-relevant genes with extreme polymorphism and very strong long-range linkage disequilibrium, which complicates the determination of the exact genes and alleles that are responsible for signals of disease association in the region^62^.

### Genetic correlation analysis

In genetic correlation analysis, for endometriosis, the GWAS meta-analysis results from the International Endometriosis Genome Consortium (IEGC) were used including 52,350 cases and 504,157 controls from 20 GWAS studies excluding UKBB to prevent overlapping study population^16^. Then genetic correlation analysis was conducted between endometriosis and immunological diseases GWAS meta-analysis results via linkage disequilibrium score regression (LDSC) analysis^17^, ^18^. The LD-score was calculated using software available at (http://github.com/bulik/ldsc), which was based on the 1000 Genomes European population and estimated within 1-cM windows, the significance threshold was set as p-value=0.00625 to account for multiple testing of eight immunological diseases.

### Mendelian randomisation analysis

The potential causal relationship between endometriosis, as exposure, and those genetically correlated immunological disease, as outcome, were investigated by two-sample Mendelian randomisation (MR) using the TwoSampleMR software^63^. As instrumental variables (IVs) we utilised the 39 established genome-wide significant LD-independent lead autosomal SNPs associated with endometriosis to assess whether endometriosis causally affects those genetic correlation immune conditions namely, osteoarthritis, rheumatoid arthritis or multiple sclerosis. Inverse variance weighted MR (MR-IVW) was applied as the initial method to detect causal effect^64^. As sensitivity analysis, other two-sample MR methods including weighted median MR^65^ and MR-Egger regression^66^ were implemented in case the assumption of valid IVs was violated. Weighted median MR was shown to have lower Type 1 error rates than the inverse-variance weighted method in a simulation analysis^65^. MR Egger provides a sensitivity analysis to detect evidence of heterogeneity and pleiotropy of IVs^66^. To detect IVs with effect of heterogeneity and pleiotropy, MR PRESSO was applied to identify outliers^67^. Also, scatter plots^68^ were generated to present the SNP-outcome association estimates versus the SNP-exposure associations in investigating the causal relationship using the MR models, including IVW, weighted median MR and MR-Egger regression^67^.

Furthermore, the strength of the 39 IVs used in this analysis was evaluated by calculating R-squared statistics using the “add_rsq()” function in the TwoSampleMR software and the total R-squared statistics for all 39 IVs is 0.298%. F statistics were calculated for all 39 IVs (a sum of Z statistics for each SNP squared) as 1656.30. Although the F statistics is relatively large for the 39 IVs, given a low R^2^ statistics for the 39 IVs used in the MR analysis, the set of IVs used for the MR analysis in this study is limited in power to assess if endometriosis is causal to certain immunological diseases.

### Multi-trait analysis of GWAS (MTAG)

The input files for MTAG are the GWAS meta-analysis summary results files which were pre- processed by filtering out: 1) SNPs with MAF =< 1%, or with a MAF differences >=20% among datasets; 2) restricting all analyses to a common set of SNPs present among datasets; 3) multiple SNPs that were mapped to an identical chromosomal position among datasets; 4) SNPs with conflicting alleles among datasets. Z scores (log(OR/SE)) were computed for all SNPs. After variant filtering, a total of 3,873,419 common SNPs between endometriosis, osteoarthritis, rheumatoid arthritis, and multiple sclerosis were included in the MTAG analysis. MTAG is a generalization of the standard inverse variance weighted meta-analysis framework. Here endometriosis, osteoarthritis, rheumatoid arthritis and multiple sclerosis pre-processed GWAS summary statistics were included in a single MTAG analysis. Within MTAG, bi-variate LD score regression is employed to account for possibly unknown sample overlap between GWAS results of different traits. In the results, MTAG outputs trait-specific effects estimated for each SNP, and the resulting p-value can be interpreted and used like those in single-trait GWAS^19^.

### Identification of shared genetic variants and their functional annotation and pathway enrichments

For each disease, genome-wide significant lead SNPs were identified based on (1) achieving a genome-wide significant P-value (P<5×10^−8^), (2) being 500kb distant from each other and (3) being independent (r^2^<0.1). Then, the genome-wide significant lead SNPs associated with respective diseases that sit within 1Mb were identified and LD between them was checked. If the LD between lead SNPs of respective disease was r^2^>=0.5, they were considered shared loci between those diseases. The identified shared lead SNPs were looked-up in (1) Genotype-Tissue Expression (GTEx) portal to identify whether they are eQTLs for genes across 49 human tissues from 838 donors with 15,201 samples^69^, and (2) eQTLGen to identify blood eQTLs from 31,684 individuals^70^. Pathway enrichment analysis was conducted in FUMA based on MTAG results for endometriosis, osteoarthritis, rheumatoid arthritis, and multiple sclerosis where pathways were limited to canonical pathways^71^.

## Data availability

The GWAS meta-analyses for immunological conditions made use of data from the UK Biobank (Application Number 9637) and publicly available GWAS summary statistics for immunological conditions [expand on sources]. GWAS data for endometriosis was based on the latest analyses of International Endogene Consortium: summary statistics excluding 23andMe data is available from EBI GWAS Catalog Study Accession GCST90205183; endometriosis GWAS summary statistics from 23andMe, Inc. were made available under a data use agreement that protects participant privacy. Please contact dataset-request@23andme. com or visit research.23andMe.com/collaborate for more information and to apply to access the data.

## Author Contributions

**Carried out data analysis:** N.S., N.R.

**Drafted the manuscript:** N.R., K.T.Z.

**Supervised the phenotypic data analysis:** K.T.Z., C.B., N.R., S.A.M., H.R.H., J.K., H.F.

**Supervised the genetic data analysis:** K.T.Z., C.B., N.R., A.P.M., C.C., J.K., H.F.

All authors contributed and discussed the results and commented on the final version of the manuscript.

## Acknowledgements

We thank all the UK Biobank and 23andMe participants. Part of this research was conducted using the UK Biobank Resource under Application Number 9637. N.R. was supported by a grant from Wellbeing of Women UK (RG2031) and the EU Horizon 2020 funded project FEMaLe (101017562).

A.P.M. was supported in part by Versus Arthritis (grant 21754). H.F. was supported by National Natural Science Foundation of China (grant 32170663). N.R., S.A.M and K.T.Z. were supported in part by a grant from CDMRP DoD PRMRP (W81XWH-20-PRMRP-IIRA). S.A.M. and K.T.Z. gratefully acknowledge funding provided by the Nezhat Family Foundation on behalf of Worldwide EndoMarch to their research programmes.

## References

1. Zondervan, K.T., Becker, C.M. & Missmer, S.A. Endometriosis. N Engl J Med 382, 1244–1256 (2020).

2. Sampson, J.A. Metastatic or Embolic Endometriosis, due to the Menstrual Dissemination of Endometrial Tissue into the Venous Circulation. Am J Pathol 3, 93–110 43 (1927).

3. Halme, J., Hammond, M.G., Hulka, J.F., Raj, S.G. & Talbert, L.M. Retrograde menstruation in healthy women and in patients with endometriosis. Obstet Gynecol 64, 151–4 (1984).

4. Cramer, D.W. & Missmer, S.A. The epidemiology of endometriosis. Ann N Y Acad Sci 955, 11–22; discussion 34-6, 396-406 (2002).

5. Bulun, S.E. et al. Role of estrogen receptor-beta in endometriosis. Semin Reprod Med 30, 39–45 (2012).

6. Han, S.J. et al. Estrogen Receptor beta Modulates Apoptosis Complexes and the Inflammasome to Drive the Pathogenesis of Endometriosis. Cell 163, 960–74 (2015).

7. Symons, L.K. et al. The Immunopathophysiology of Endometriosis. Trends Mol Med 24, 748–762 (2018).

8. Zondervan, K.T. et al. Endometriosis. Nat Rev Dis Primers 4, 9 (2018).

9. Matarese, G., De Placido, G., Nikas, Y. & Alviggi, C. Pathogenesis of endometriosis: natural immunity dysfunction or autoimmune disease? Trends Mol Med 9, 223–8 (2003).

10. Gleicher, N., el-Roeiy, A., Confino, E. & Friberg, J. Is endometriosis an autoimmune disease? Obstet Gynecol 70, 115–22 (1987).

11. Eisenberg, V.H., Zolti, M. & Soriano, D. Is there an association between autoimmunity and endometriosis? Autoimmun Rev 11, 806–14 (2012).

12. Laudanski, P. et al. Autoantibody screening of plasma and peritoneal fluid of patients with endometriosis. Hum Reprod 38, 629–643 (2023).

13. https://www.autoimmuneinstitute.org/diseases_list/endometriosis/.

14. Shigesi, N. et al. The association between endometriosis and autoimmune diseases: a systematic review and meta-analysis. Hum Reprod Update 25, 486–503 (2019).

15. McGonagle, D. & McDermott, M.F. A proposed classification of the immunological diseases. PLoS Med 3, e297 (2006).

16. Rahmioglu, N. et al. The genetic basis of endometriosis and comorbidity with other pain and inflammatory conditions. Nat Genet 55, 423–436 (2023).

17. Bulik-Sullivan, B. et al. An atlas of genetic correlations across human diseases and traits. Nat Genet 47, 1236–41 (2015).

18. Bulik-Sullivan, B.K. et al. LD Score regression distinguishes confounding from polygenicity in genome-wide association studies. Nat Genet 47, 291–5 (2015).

19. Turley, P. et al. Multi-trait analysis of genome-wide association summary statistics using MTAG. Nat Genet 50, 229–237 (2018).

20. Manco, G., Porzio, E. & Carusone, T.M. Human Paraoxonase-2 (PON2): Protein Functions and Modulation. Antioxidants (Basel) 10(2021).

21. Shin, S.H. et al. Arrest defective 1 regulates the oxidative stress response in human cells and mice by acetylating methionine sulfoxide reductase A. Cell Death Dis 5, e1490 (2014).

22. Ichikawa-Tomikawa, N., Sugimoto, K., Kashiwagi, K. & Chiba, H. The Src-Family Kinases SRC and BLK Contribute to the CLDN6-Adhesion Signaling. Cells 12(2023).

23. Wang, H. et al. ZAP-70: an essential kinase in T-cell signaling. Cold Spring Harb Perspect Biol 2, a002279 (2010).

24. Li, C. et al. Transmembrane Protein 214 (TMEM214) mediates endoplasmic reticulum stress-induced caspase 4 enzyme activation and apoptosis. J Biol Chem 288, 17908–17 (2013).

25. Stelzer, G. et al. The GeneCards Suite: From Gene Data Mining to Disease Genome Sequence Analyses. Curr Protoc Bioinformatics 54, 1 30 1–1 30 33 (2016).

26. Dyson, M.T. et al. Genome-wide DNA methylation analysis predicts an epigenetic switch for GATA factor expression in endometriosis. PLoS Genet 10, e1004158 (2014).

27. McKinnon, B.D., Kocbek, V., Nirgianakis, K., Bersinger, N.A. & Mueller, M.D. Kinase signalling pathways in endometriosis: potential targets for non-hormonal therapeutics. Hum Reprod Update 22, 382–403 (2016).

28. Milacic, M. et al. The Reactome Pathway Knowledgebase 2024. Nucleic Acids Res 52, D672–D678 (2024).

29. Shafrir, A.L. et al. Risk for and consequences of endometriosis: A critical epidemiologic review. Best Pract Res Clin Obstet Gynaecol 51, 1–15 (2018).

30. Zondervan, K.T., Cardon, L.R. & Kennedy, S.H. What makes a good case-control study? Design issues for complex traits such as endometriosis. Hum Reprod 17, 1415–23 (2002).

31. Harris, H.R. et al. Endometriosis, Psoriasis, and Psoriatic Arthritis: A Prospective Cohort Study. Am J Epidemiol 191, 1050–1060 (2022).

32. Harris, H.R. et al. Endometriosis and the risks of systemic lupus erythematosus and rheumatoid arthritis in the Nurses’ Health Study II. Ann Rheum Dis 75, 1279–84 (2016).

33. Kraft, P., Chen, H. & Lindstrom, S. The Use Of Genetic Correlation And Mendelian Randomization Studies To Increase Our Understanding of Relationships Between Complex Traits. Curr Epidemiol Rep 7, 104–112 (2020).

34. Burgess, S. Sample size and power calculations in Mendelian randomization with a single instrumental variable and a binary outcome. Int J Epidemiol 43, 922–9 (2014).

35. Shafrir, A.L. et al. Co-occurrence of immune-mediated conditions and endometriosis among adolescents and adult women. Am J Reprod Immunol 86, e13404 (2021).

36. Ke, J., Ye, J., Li, M. & Zhu, Z. The Role of Matrix Metalloproteinases in Endometriosis: A Potential Target. Biomolecules 11(2021).

37. Liu, Y. et al. Increased Serum Matrix Metalloproteinase-9 Levels are Associated with Anti- Jo1 but not Anti-MDA5 in Myositis Patients. Aging Dis 10, 746–755 (2019).

38. Ram, M., Sherer, Y. & Shoenfeld, Y. Matrix metalloproteinase-9 and autoimmune diseases. J Clin Immunol 26, 299–307 (2006).

39. Du, F. et al. Inflammatory Th17 Cells Express Integrin alphavbeta3 for Pathogenic Function. Cell Rep 16, 1339–1351 (2016).

40. Jang, Y., Kim, M. & Hwang, S.W. Molecular mechanisms underlying the actions of arachidonic acid-derived prostaglandins on peripheral nociception. J Neuroinflammation 17, 30 (2020).

41. Migliore, A. & Procopio, S. Effectiveness and utility of hyaluronic acid in osteoarthritis. Clin Cases Miner Bone Metab 12, 31–3 (2015).

42. Kobayashi, T., Chanmee, T. & Itano, N. Hyaluronan: Metabolism and Function. Biomolecules 10(2020).

43. Yu, P.H., Chou, P.Y., Li, W.N., Tsai, S.J. & Wu, M.H. The pro-inflammatory and anti-inflammatory role of hyaluronic acid in endometriosis. Taiwan J Obstet Gynecol 60, 711–717 (2021).

44. Simons, K. & Toomre, D. Lipid rafts and signal transduction. Nat Rev Mol Cell Biol 1, 31–9 (2000).

45. Robinson, G., Pineda-Torra, I., Ciurtin, C. & Jury, E.C. Lipid metabolism in autoimmune rheumatic disease: implications for modern and conventional therapies. J Clin Invest 132(2022).

46. Voskuhl, R. Sex differences in autoimmune diseases. Biol Sex Differ 2, 1 (2011).

47. Boer, C.G. et al. Deciphering osteoarthritis genetics across 826,690 individuals from 9 populations. Cell 184, 6003–6005 (2021).

48. Sudlow, C. et al. UK biobank: an open access resource for identifying the causes of a wide range of complex diseases of middle and old age. PLoS Med 12, e1001779 (2015).

49. Rahmioglu, N. et al. Genome-wide enrichment analysis between endometriosis and obesity-related traits reveals novel susceptibility loci. Hum Mol Genet 24, 1185–99 (2015).

50. Kvaskoff, M. et al. Endometriosis: a high-risk population for major chronic diseases? Hum Reprod Update 21, 500–16 (2015).

51. Atsma, F., Bartelink, M.L., Grobbee, D.E. & van der Schouw, Y.T. Postmenopausal status and early menopause as independent risk factors for cardiovascular disease: a meta-analysis. Menopause 13, 265–79 (2006).

52. Surrey, E., Soliman, A.M., Trenz, H., Blauer-Peterson, C. & Sluis, A. Impact of Endometriosis Diagnostic Delays on Healthcare Resource Utilization and Costs. Adv Ther 37, 1087–1099 (2020).

53. Loh, P.R. et al. Efficient Bayesian mixed-model analysis increases association power in large cohorts. Nat Genet 47, 284–90 (2015).

54. de Lange, K.M. et al. Genome-wide association study implicates immune activation of multiple integrin genes in inflammatory bowel disease. Nat Genet 49, 256–261 (2017).

55. International Genetics of Ankylosing Spondylitis, C., et al. Identification of multiple risk variants for ankylosing spondylitis through high-density genotyping of immune-related loci. Nat Genet 45, 730–8 (2013).

56. International Multiple Sclerosis Genetics, C. Multiple sclerosis genomic map implicates peripheral immune cells and microglia in susceptibility. Science 365(2019).

57. Okada, Y. et al. Genetics of rheumatoid arthritis contributes to biology and drug discovery. Nature 506, 376–81 (2014).

58. Ricano-Ponce, I. et al. Immunochip meta-analysis in European and Argentinian populations identifies two novel genetic loci associated with celiac disease. Eur J Hum Genet 28, 313–323 (2020).

59. Stuart, P.E., et al. Transethnic analysis of psoriasis susceptibility in South Asians and Europeans enhances fine-mapping in the MHC and genomewide. HGG Adv 3(2022).

60. Wang, Y.F. et al. Identification of 38 novel loci for systemic lupus erythematosus and genetic heterogeneity between ancestral groups. Nat Commun 12, 772 (2021).

61. Willer, C.J., Li, Y. & Abecasis, G.R. METAL: fast and efficient meta-analysis of genomewide association scans. Bioinformatics 26, 2190–1 (2010).

62. Trowsdale, J. & Knight, J.C. Major histocompatibility complex genomics and human disease. Annu Rev Genomics Hum Genet 14, 301–23 (2013).

63. Hemani, G. et al. The MR-Base platform supports systematic causal inference across the human phenome. Elife 7(2018).

64. Burgess, S., Butterworth, A. & Thompson, S.G. Mendelian randomization analysis with multiple genetic variants using summarized data. Genet Epidemiol 37, 658–65 (2013).

65. Bowden, J., Davey Smith, G., Haycock, P.C. & Burgess, S. Consistent Estimation in Mendelian Randomization with Some Invalid Instruments Using a Weighted Median Estimator. Genet Epidemiol 40, 304–14 (2016).

66. Bowden, J., Davey Smith, G. & Burgess, S. Mendelian randomization with invalid instruments: effect estimation and bias detection through Egger regression. Int J Epidemiol 44, 512–25 (2015).

67. Verbanck, M., Chen, C.Y., Neale, B. & Do, R. Detection of widespread horizontal pleiotropy in causal relationships inferred from Mendelian randomization between complex traits and diseases. Nat Genet 50, 693–698 (2018).

68. Bowden, J. & Holmes, M.V. Meta-analysis and Mendelian randomization: A review. Res Synth Methods 10, 486–496 (2019).

69. Consortium, G.T. et al. Genetic effects on gene expression across human tissues. Nature 550, 204–213 (2017).

70. Vosa, U. et al. Large-scale cis- and trans-eQTL analyses identify thousands of genetic loci and polygenic scores that regulate blood gene expression. Nat Genet 53, 1300–1310 (2021).

71. Watanabe, K., Taskesen, E., van Bochoven, A. & Posthuma, D. Functional mapping and annotation of genetic associations with FUMA. Nat Commun 8, 1826 (2017).

72. Dubois, P.C. et al. Multiple common variants for celiac disease influencing immune gene expression. Nat Genet 42, 295–302 (2010).

73. Tachmazidou, I. et al. Identification of new therapeutic targets for osteoarthritis through genome-wide analyses of UK Biobank data. Nat Genet 51, 230–236 (2019).

74. Bentham, J. et al. Genetic association analyses implicate aberrant regulation of innate and adaptive immunity genes in the pathogenesis of systemic lupus erythematosus. Nat Genet 47, 1457–1464 (2015).

