## Supplementary Figures 1-10 for "The Association between Endometriosis and Immunological diseases"

**Supplementary Figure 1.** Ankylosing spondylitis GWAS results. (a) Manhattan plots and (b) Q-Q plots for i. female-only and ii. sex-combined GWAS summary results.

**(a) i. Female-only UKBB GWAS**

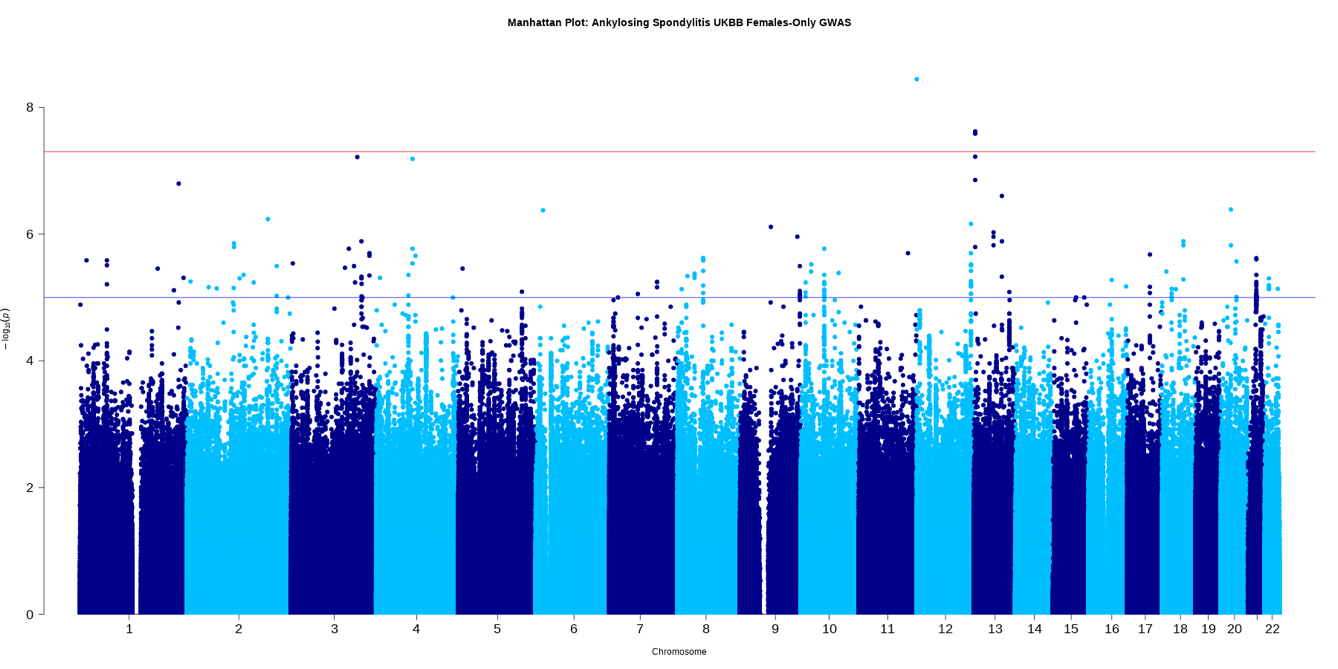

(a) **ii. Sex-combined UKBB GWAS**

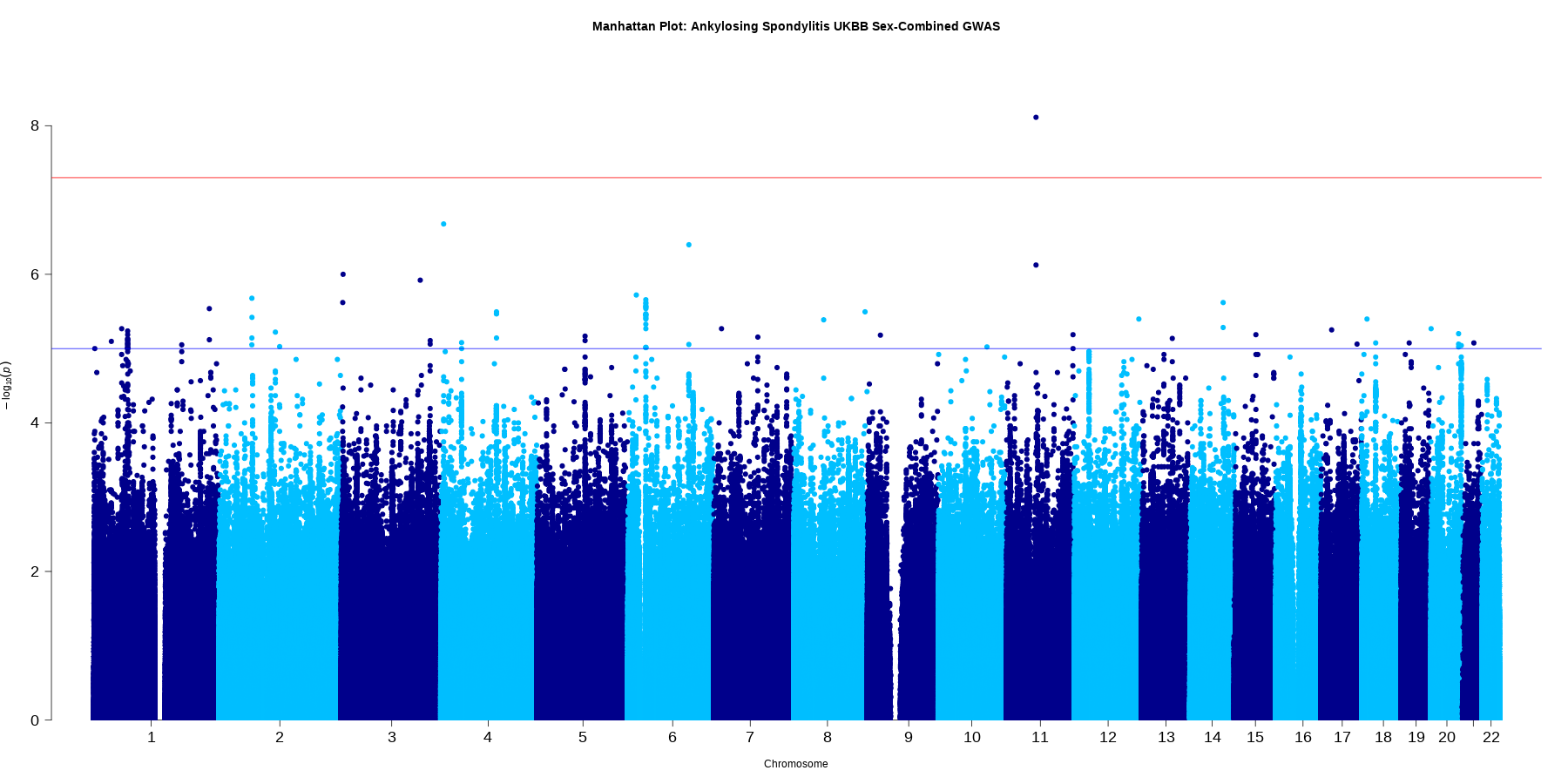

(b) **Q-Q plots for i. Female-only and ii. Sex-combined UKBB GWAS**

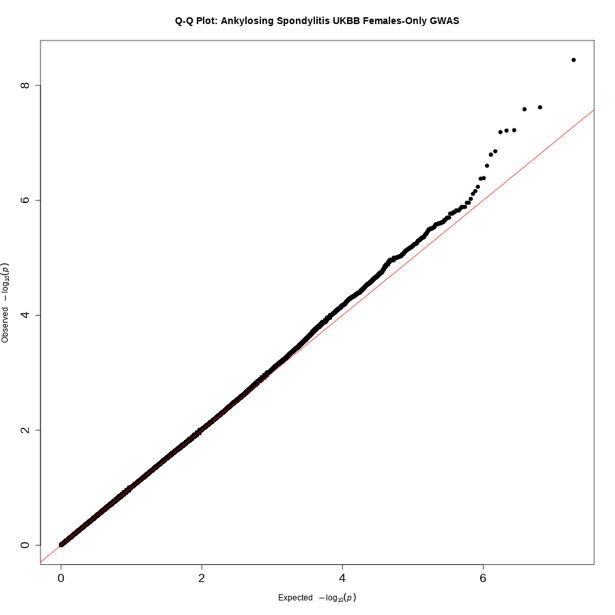

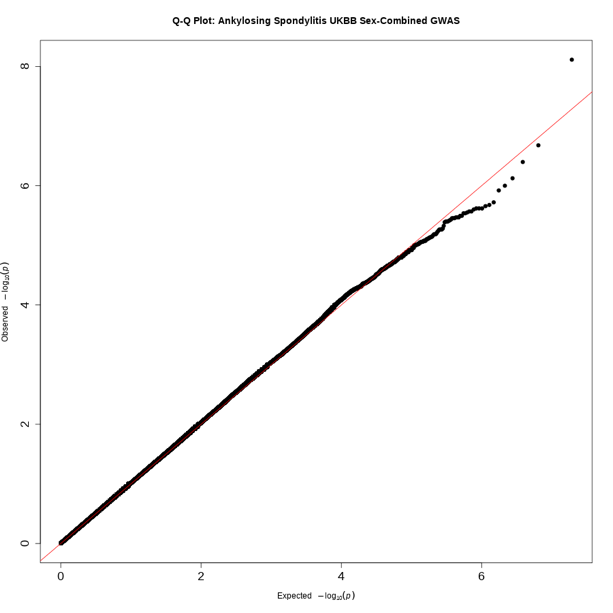

**Supplementary Figure 2.** Coeliac disease GWAS results. (a) Manhattan plots and (b) Q-Q plots for i. female-only, ii. sex-combined GWAS and iii. Meta-analysis of sex-combined summary results.

**(a) i. Female-only UKBB GWAS**

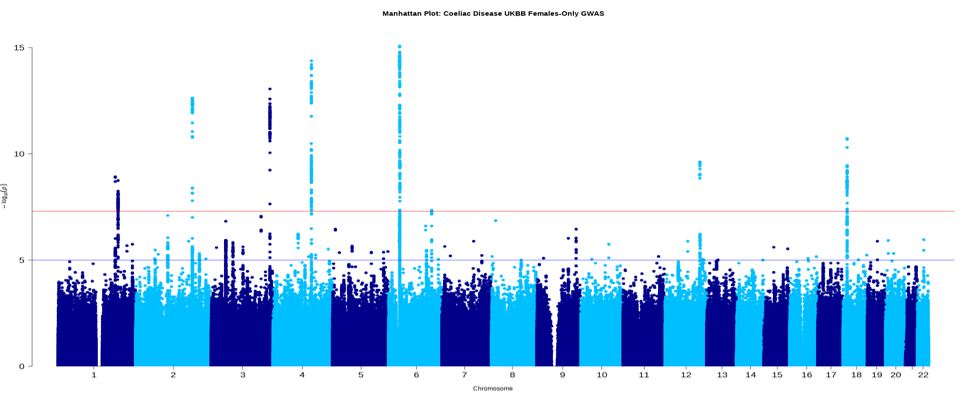

(a) **ii. Sex-combined UKBB GWAS**

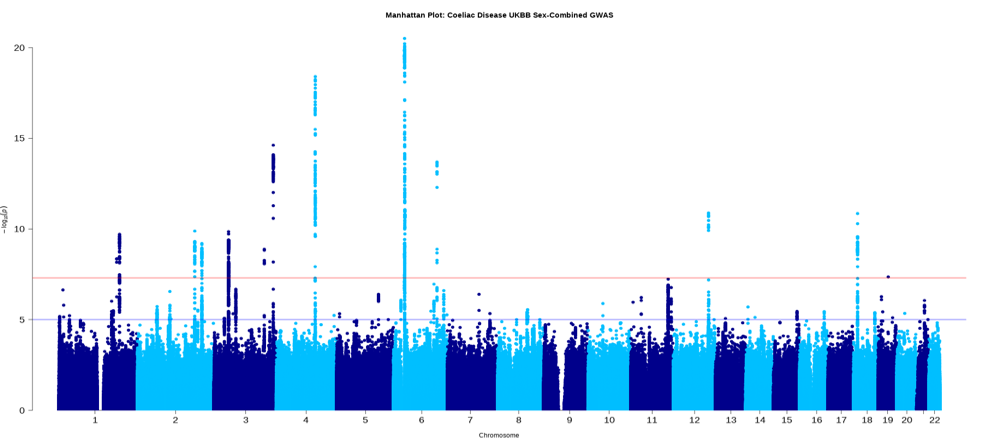

**(a) iii. Meta-analysis of sex-combined GWAS results**

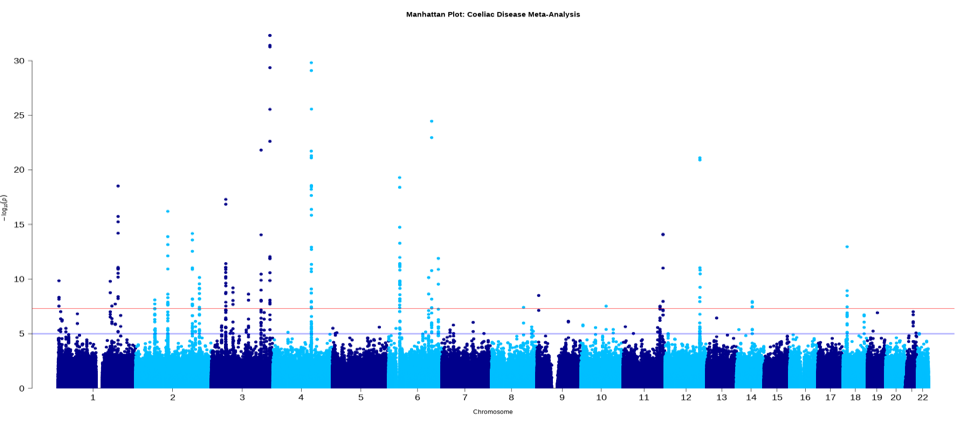

**(b) Q-Q plots for i. Female-only, ii. Sex-combined iii. Meta-analysis of sex-combined GWAS.**

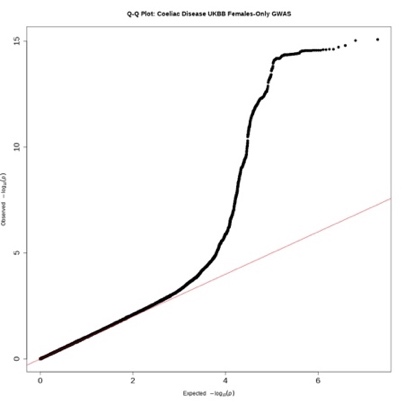

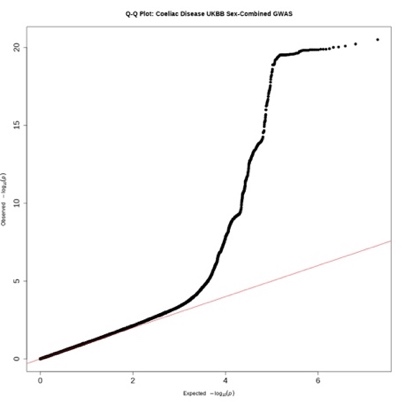

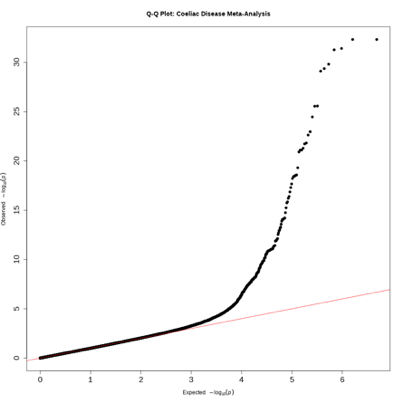

**Supplementary Figure 3.** Inflammatory bowel disease GWAS results. (a) Manhattan plots, (b) Q-Q plots for i. female-only, ii. sex-combined GWAS, iii. Meta-analysis of sex-combined summary results.

**(a) i. Female-only UKBB GWAS**

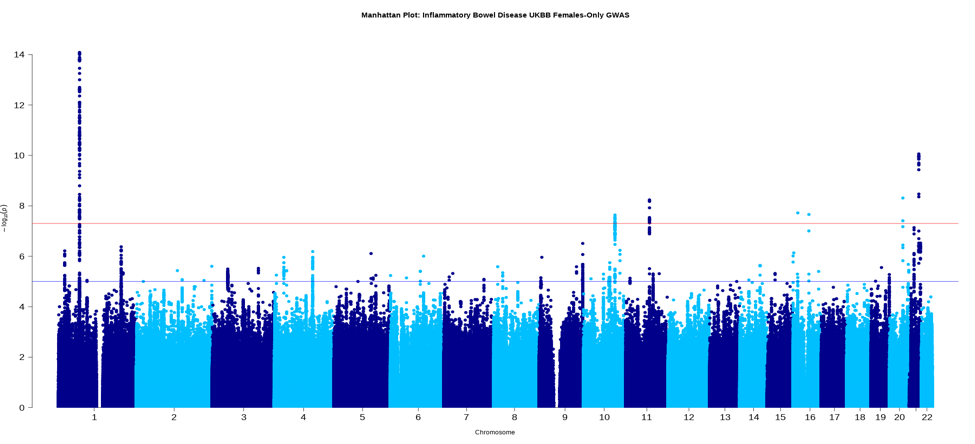

**(a) ii. Sex-combined UKBB GWAS**

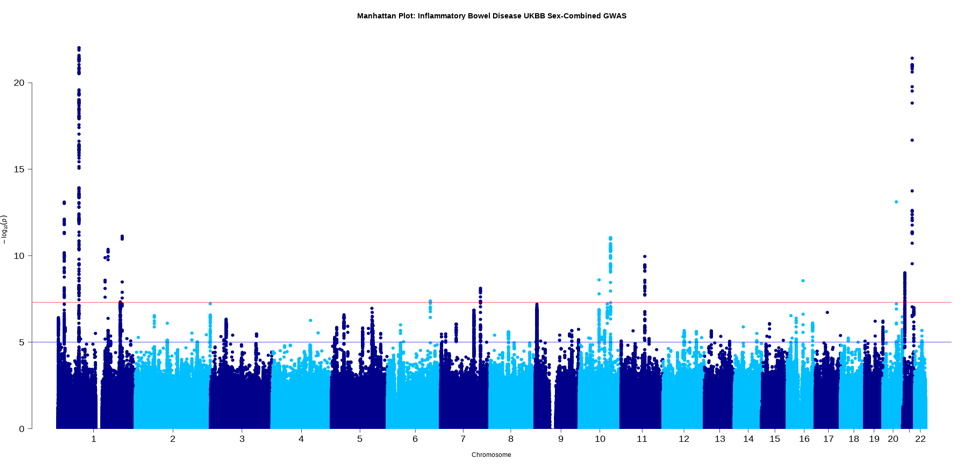

**(a) iii. Meta-analysis of sex-combined GWAS results**

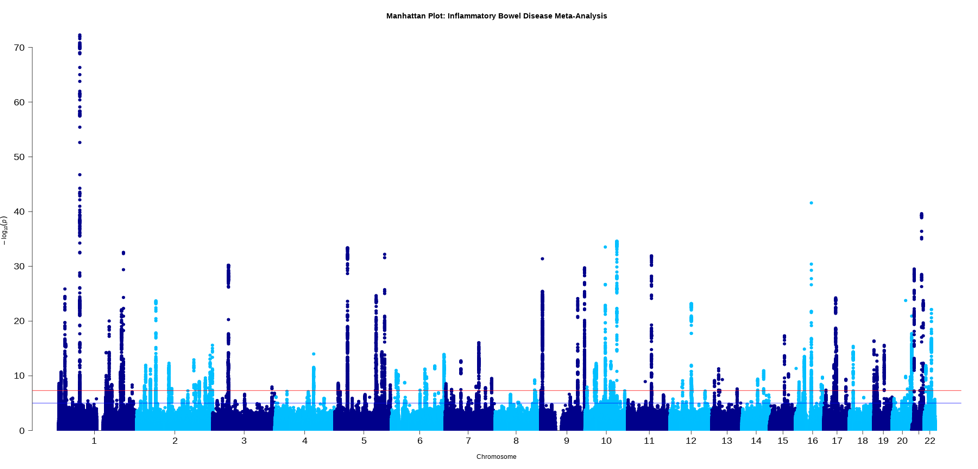

**(b) Q-Q plots for i. Female-only, ii. Sex-combined iii. Meta-analysis of sex-combined GWAS.**

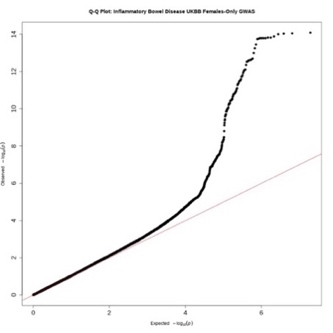

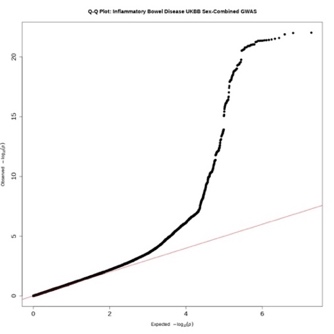

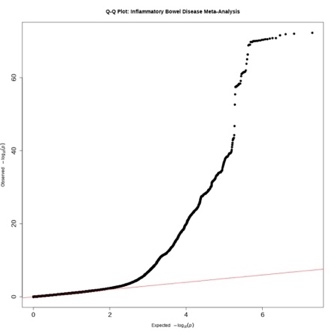

**Supplementary Figure 4.** Multiple Sclerosis GWAS results. (a) Manhattan plots, (b) Q-Q plots for i. female-only, ii. sex-combined GWAS, iii. Meta-analysis of sex-combined summary results.

**(a) i. Female-only UKBB GWAS**

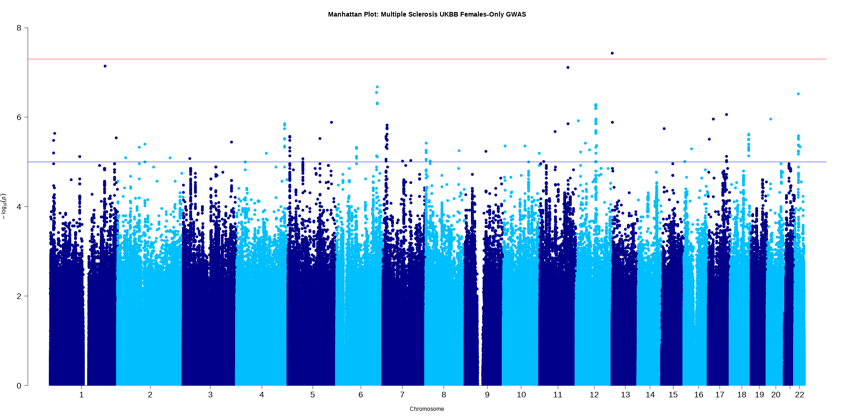

**(a) ii. Sex-combined UKBB GWAS**

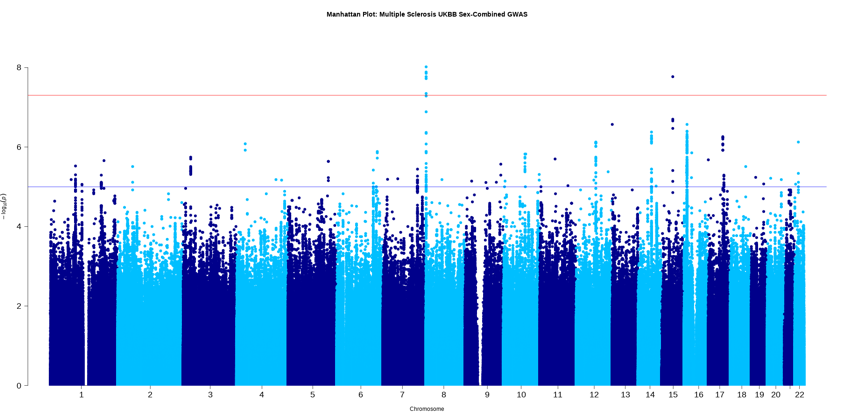

**(a) iii. Meta-analysis of sex-combined GWAS results**

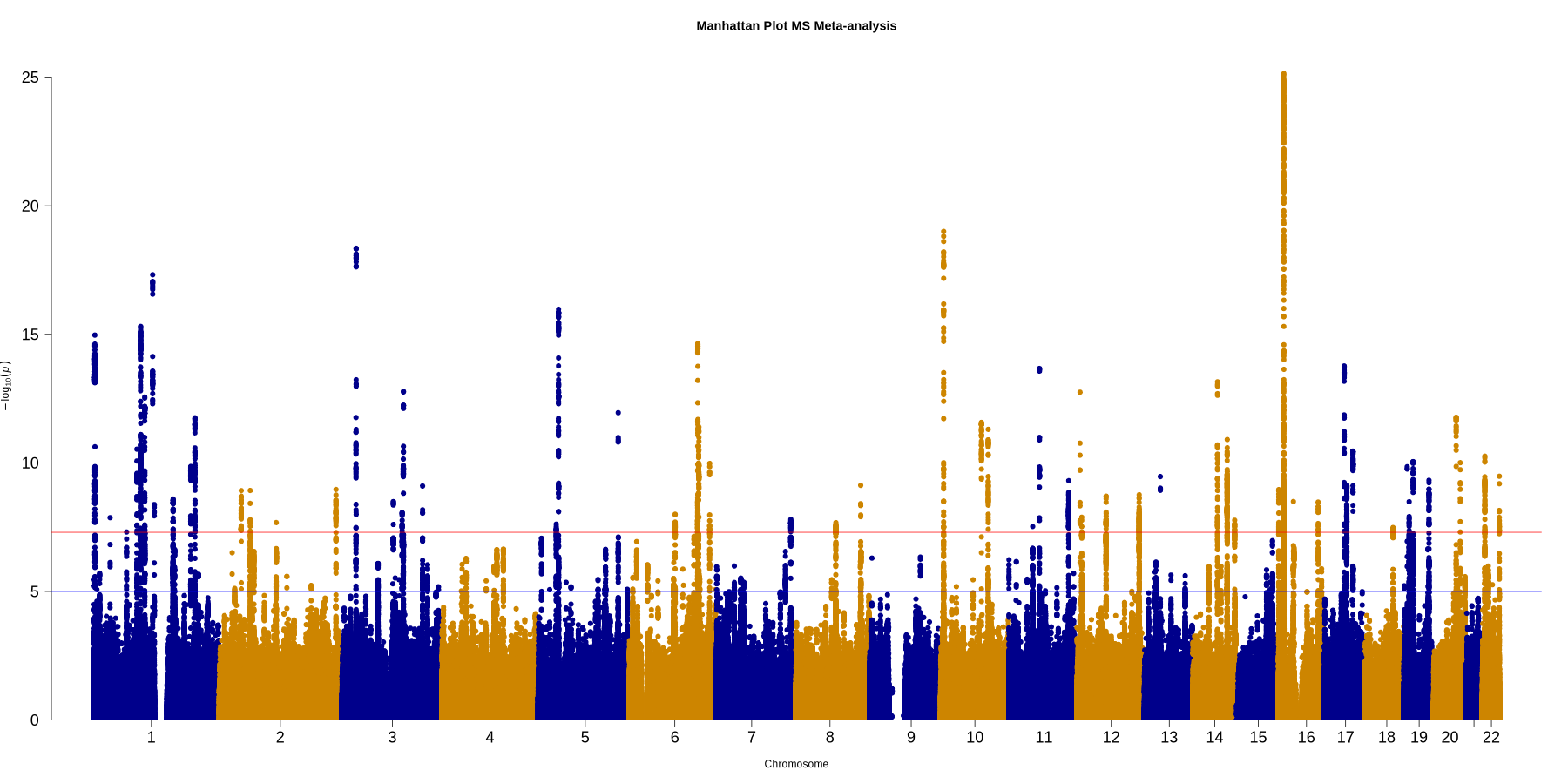

**(b) Q-Q plots for i. Female-only, ii. Sex-combined iii. Meta-analysis of sex-combined GWAS.**

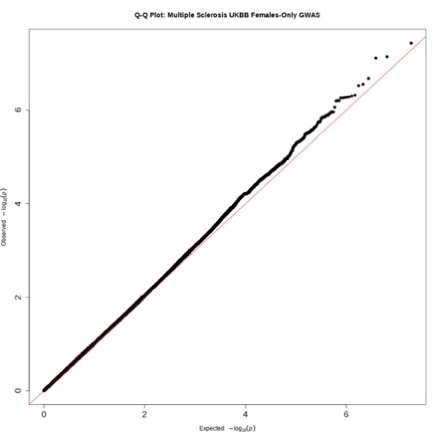

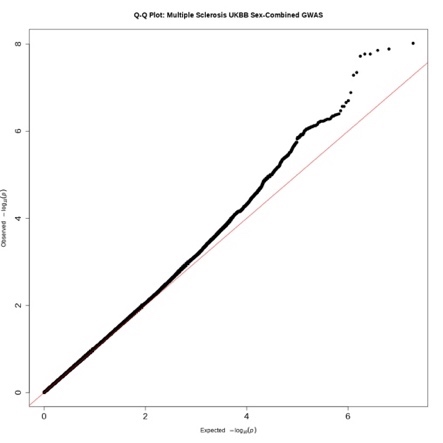

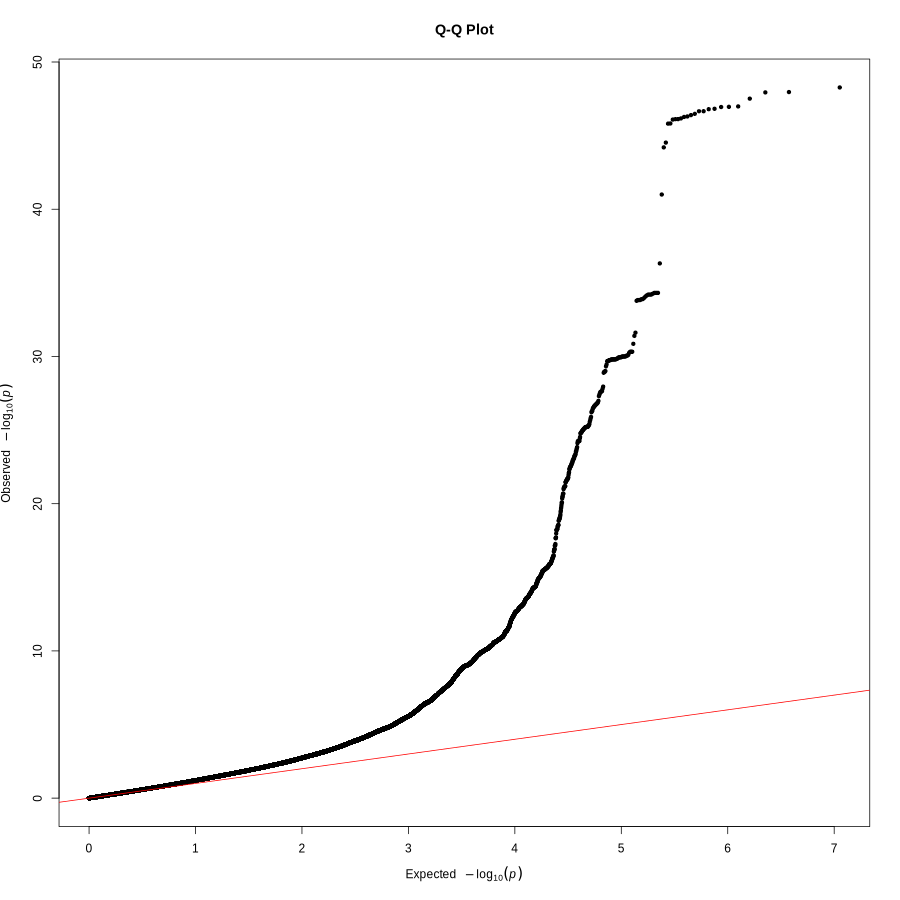

**Supplementary Figure 5.** Osteoarthritis GWAS results. (a) Manhattan plots, (b) Q-Q plots for i. female-only, ii. sex-combined GWAS, iii. Meta-analysis of sex-combined summary results.

**(a) i. Female-only UKBB GWAS**

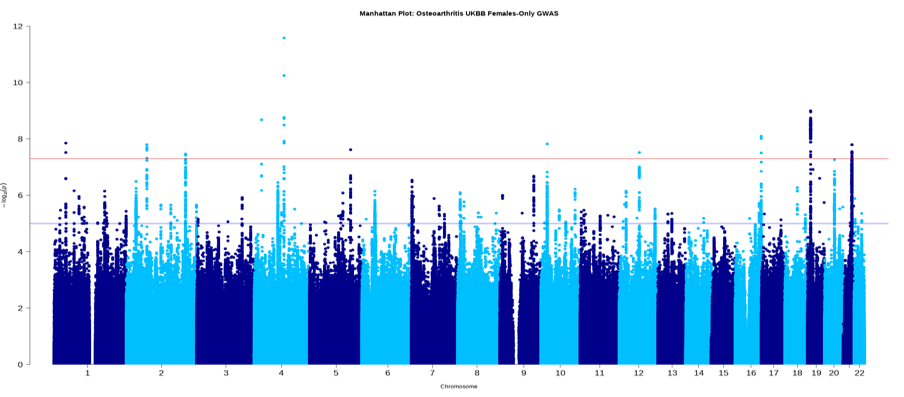

**(a) ii. Sex-combined UKBB GWAS**

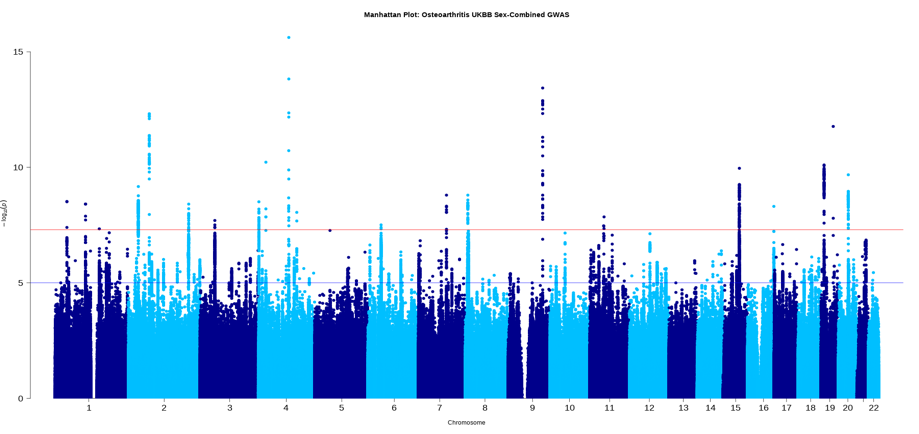

**(a) iii. Meta-analysis of sex-combined GWAS results**

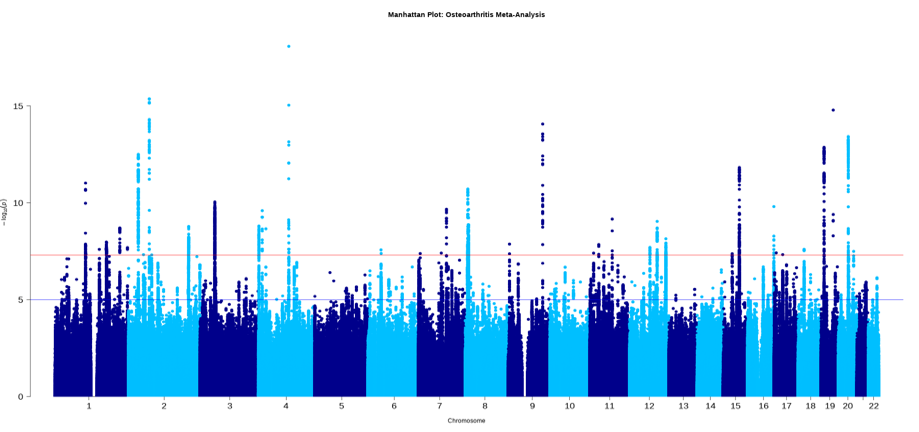

**(b) Q-Q plots for i. Female-only, ii. Sex-combined iii. Meta-analysis of sex-combined GWAS.**

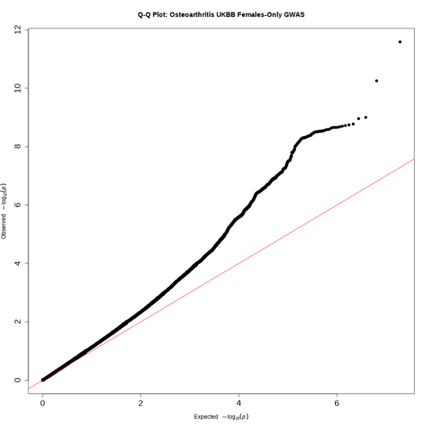

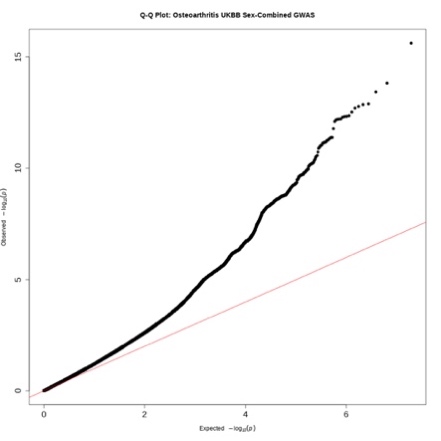

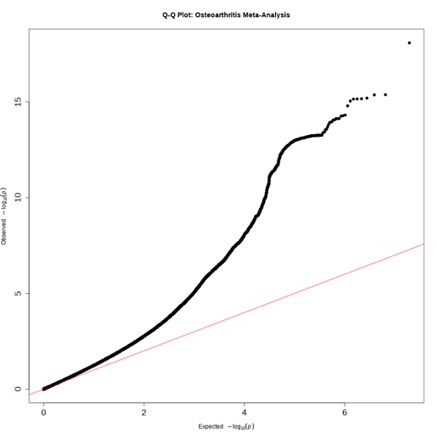

**Supplementary Figure 6.** Psoriasis GWAS results. (a) Manhattan plots, (b) Q-Q plots for i. female-only, ii. sex-combined GWAS, iii. Meta-analysis of sex-combined summary results.

**(a) i. Female-only UKBB GWAS**

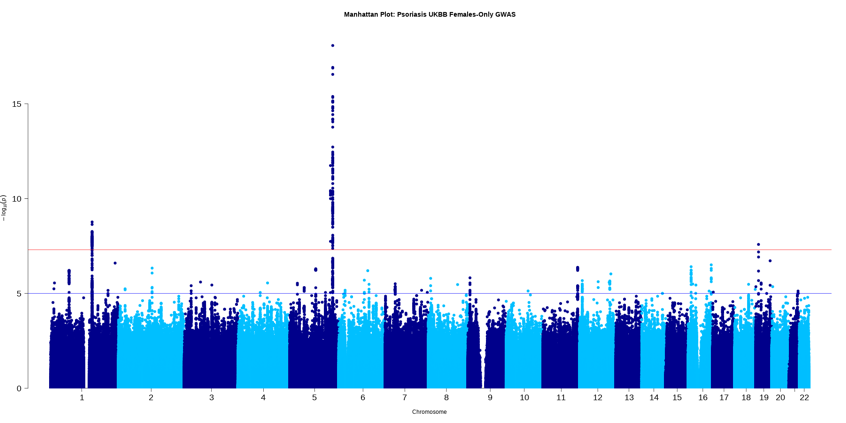

**(a) ii. Sex-combined UKBB GWAS**

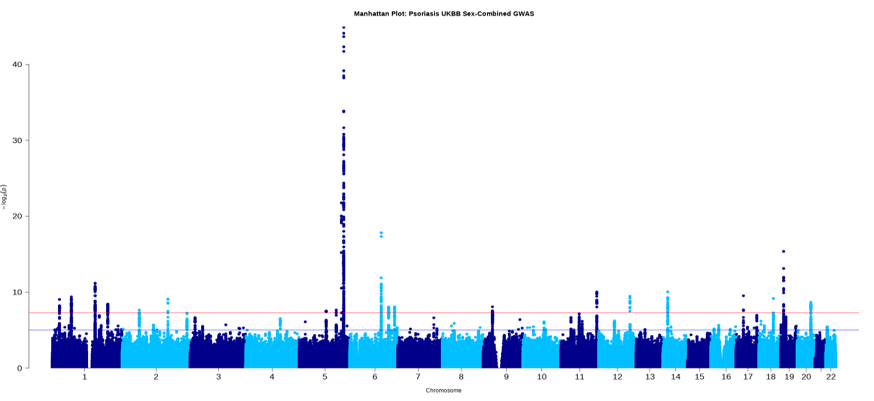

**(a) iii. Meta-analysis of sex-combined GWAS results**

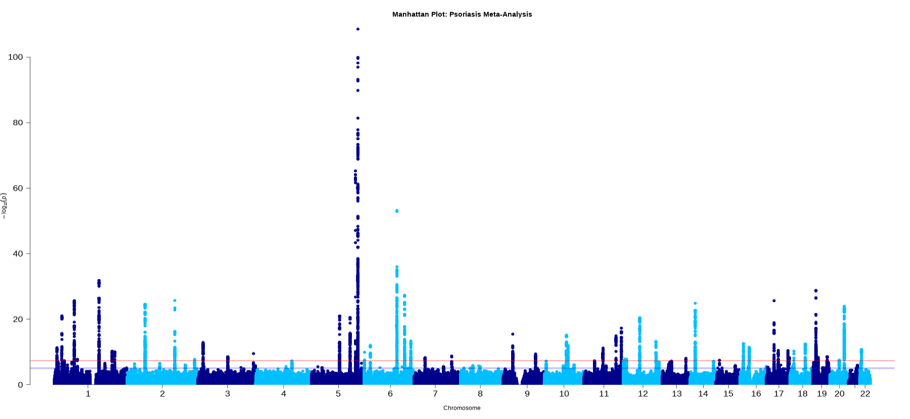

**(b) Q-Q plots for i. Female-only, ii. Sex-combined iii. Meta-analysis of sex-combined GWAS.**

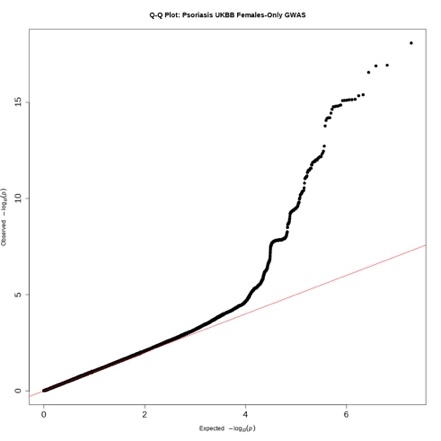

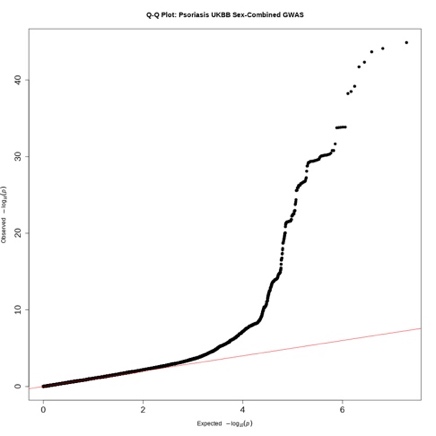

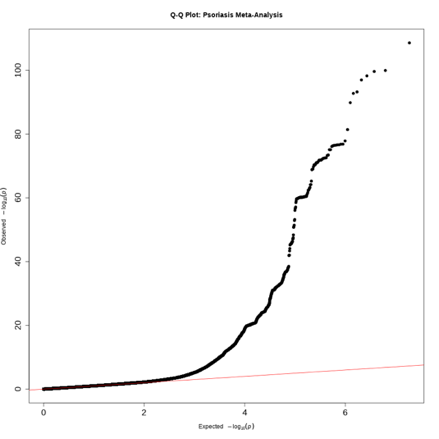

**Supplementary Figure 7.** Rheumatoid arthritis GWAS results. (a) Manhattan plots, (b) Q-Q plots for i. female-only, ii. sex-combined GWAS, iii. Meta-analysis of sex-combined summary results.

**(a) i. Female-only UKBB GWAS**

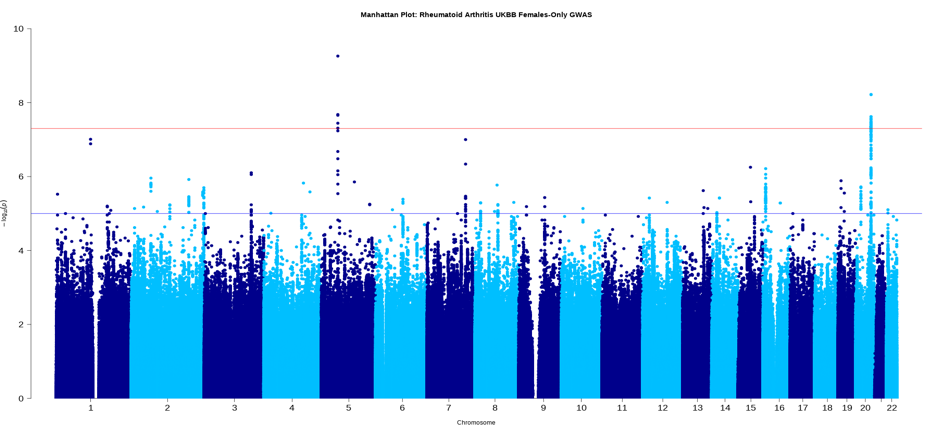

**(a) ii. Sex-combined UKBB GWAS**

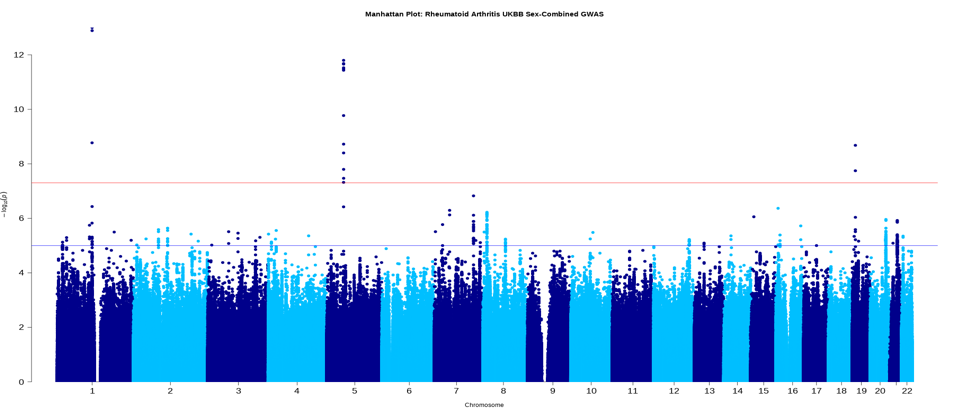

**(a) iii. Meta-analysis of sex-combined GWAS results**
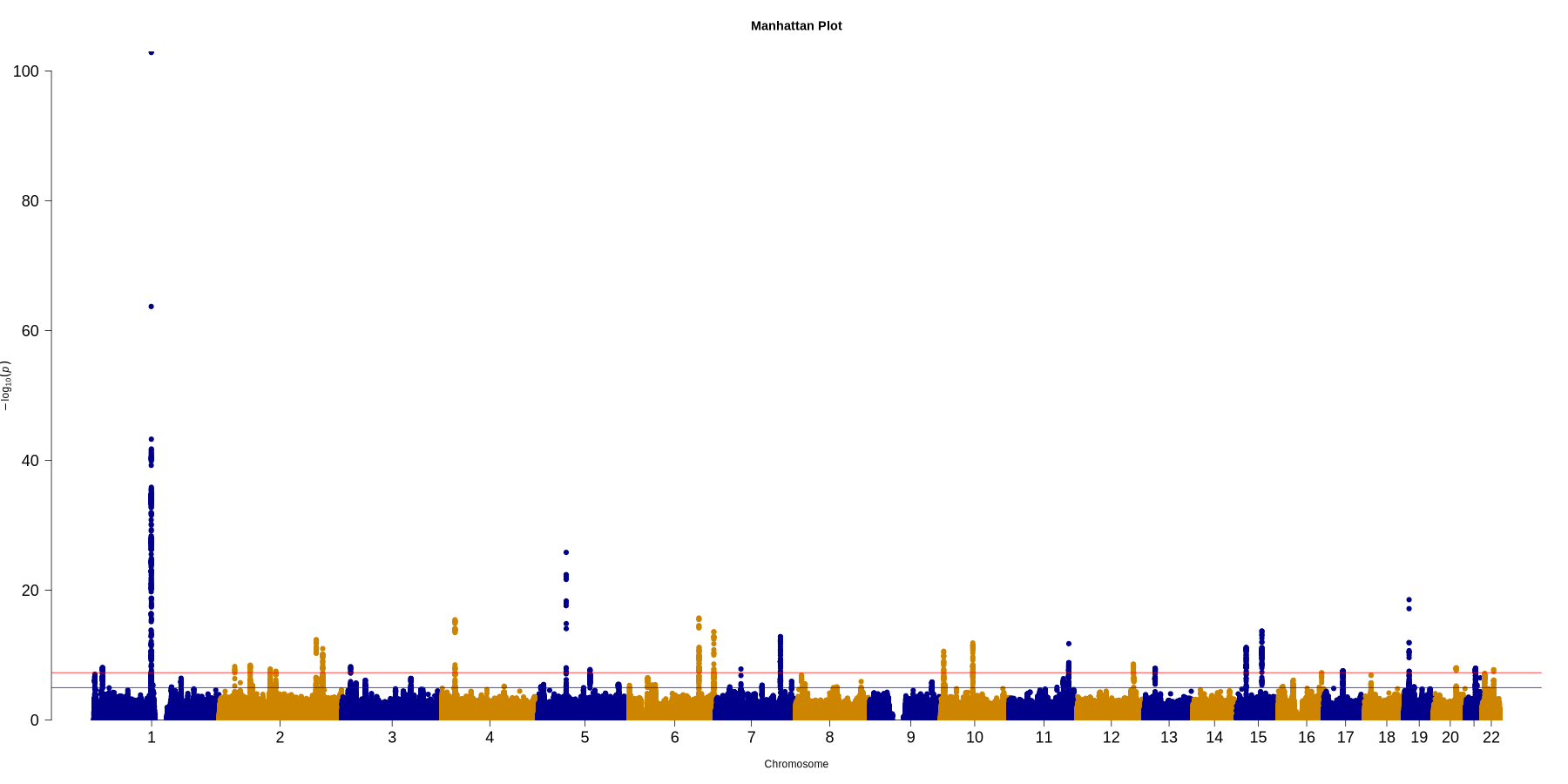

**(b) Q-Q plots for i. Female-only, ii. Sex-combined iii. Meta-analysis of sex-combined GWAS.**

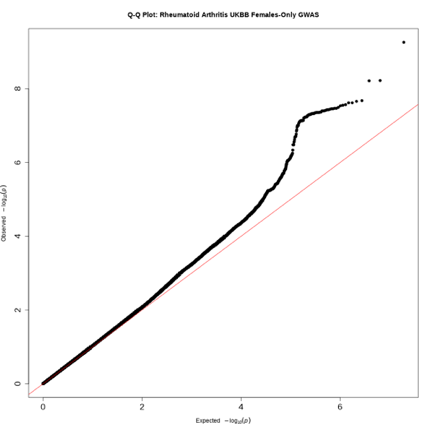

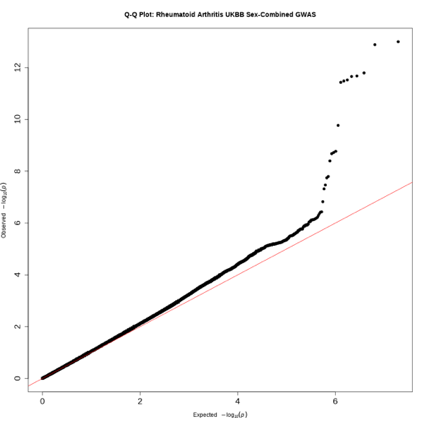

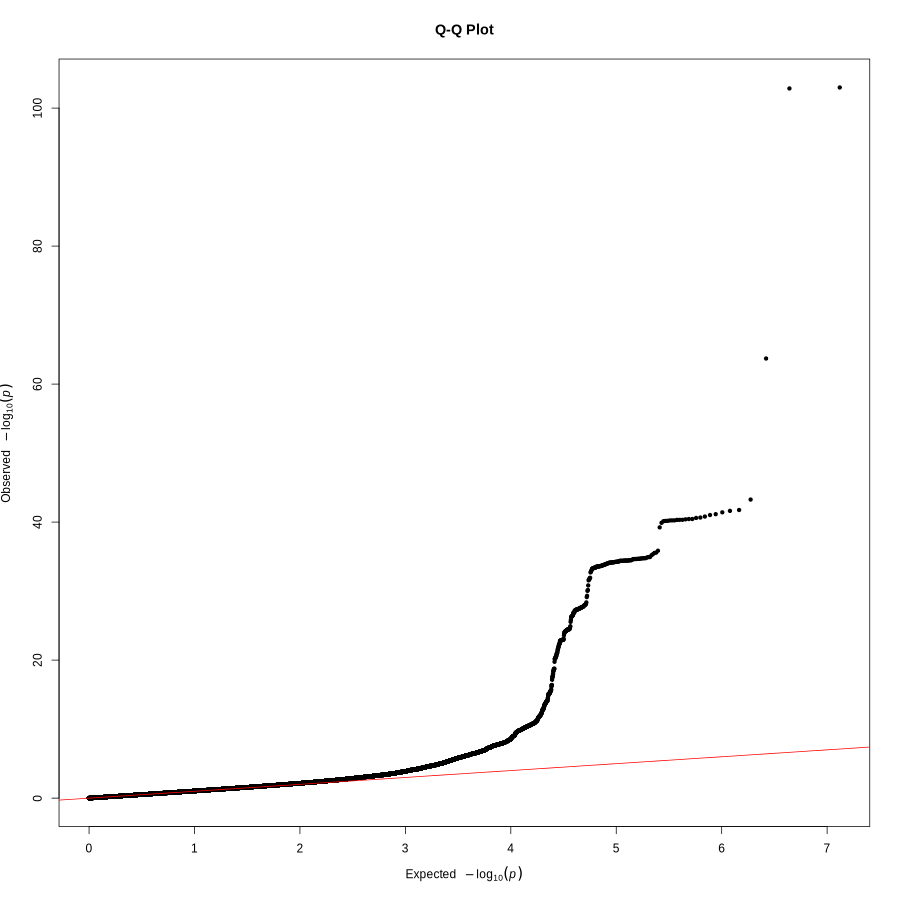

**Supplementary Figure 8.** Systemic lupus erythematosus GWAS results. (a) Manhattan plots, (b) Q-Q plots for i. female-only, ii. sex-combined, iii. Meta-analysis of sex-combined summary results.

**(a) i. Female-only UKBB GWAS**

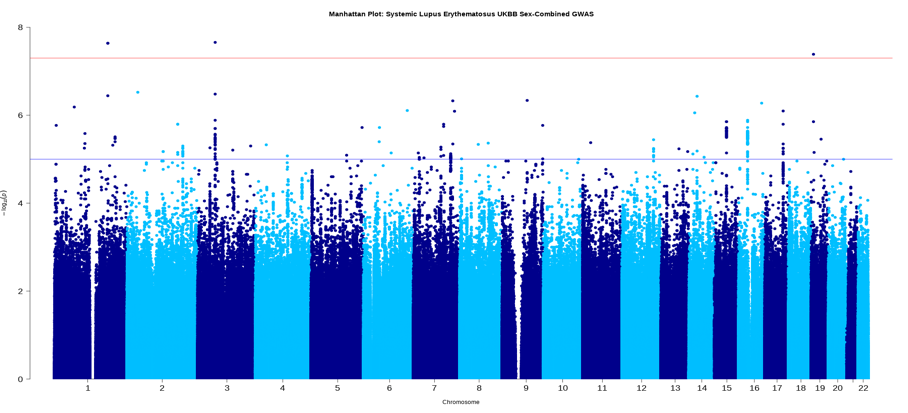

**(a) ii. Sex-combined UKBB GWAS**

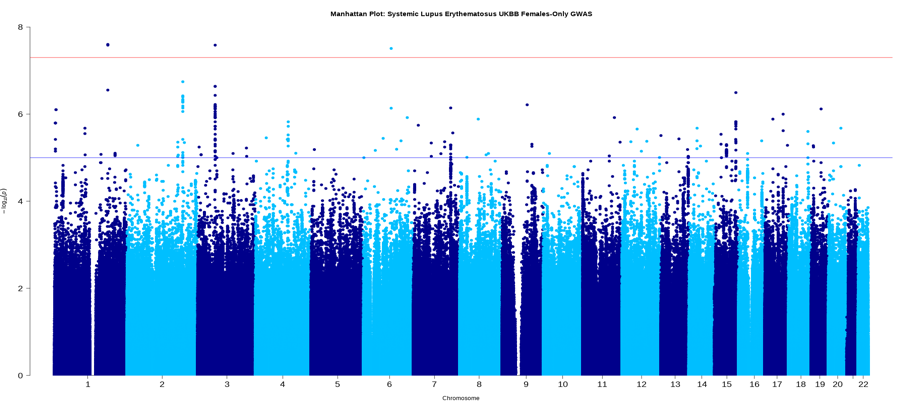

**(a) iii. Meta-analysis of sex-combined GWAS results**
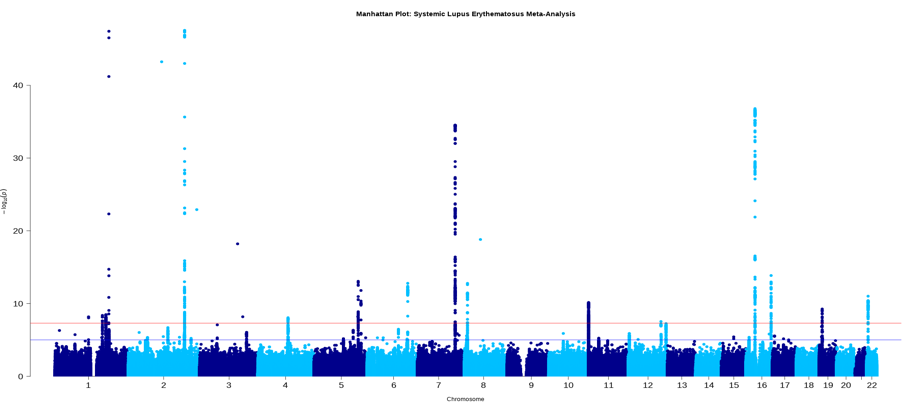

**(b) Q-Q plots for i. Female-only, ii. Sex-combined iii. Meta-analysis of sex-combined GWAS.**

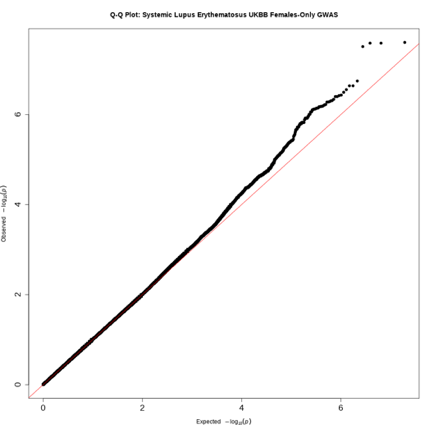

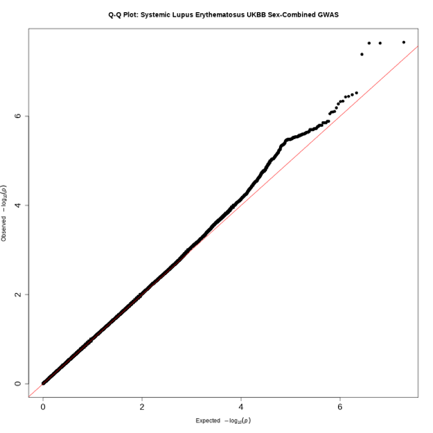

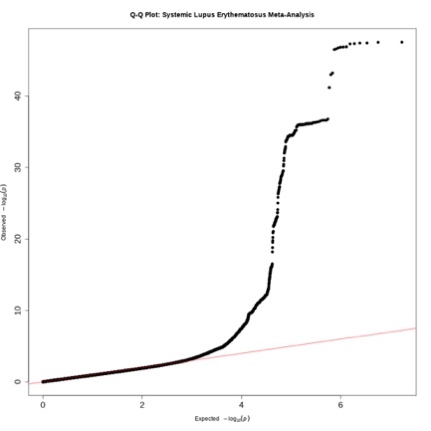

**Supplementary Figure 9.** Regional association plots for 6 novel genome-wide significant endometriosis loci from MTAG: (a) *ABHD1/*2p23.3, (b) *TMEM131/*2q11.2, (c) *XRCC4/*5q14.2, (d) *PPP1R9A/*7q21.3, (e) *XKR6/*8p23.1, (f) *TRPS1/*8p23.3. The association results are shown on the y-axis as –log_10_(P-value) and on the x-axis is the genomic location (hg 19). The top associated SNP is coloured purple and the other SNPs are coloured according to the strength of LD with the top SNP by r^2^ in the European 1000 Genomes dataset.

**(a) *ABHD1/*2p23.3**

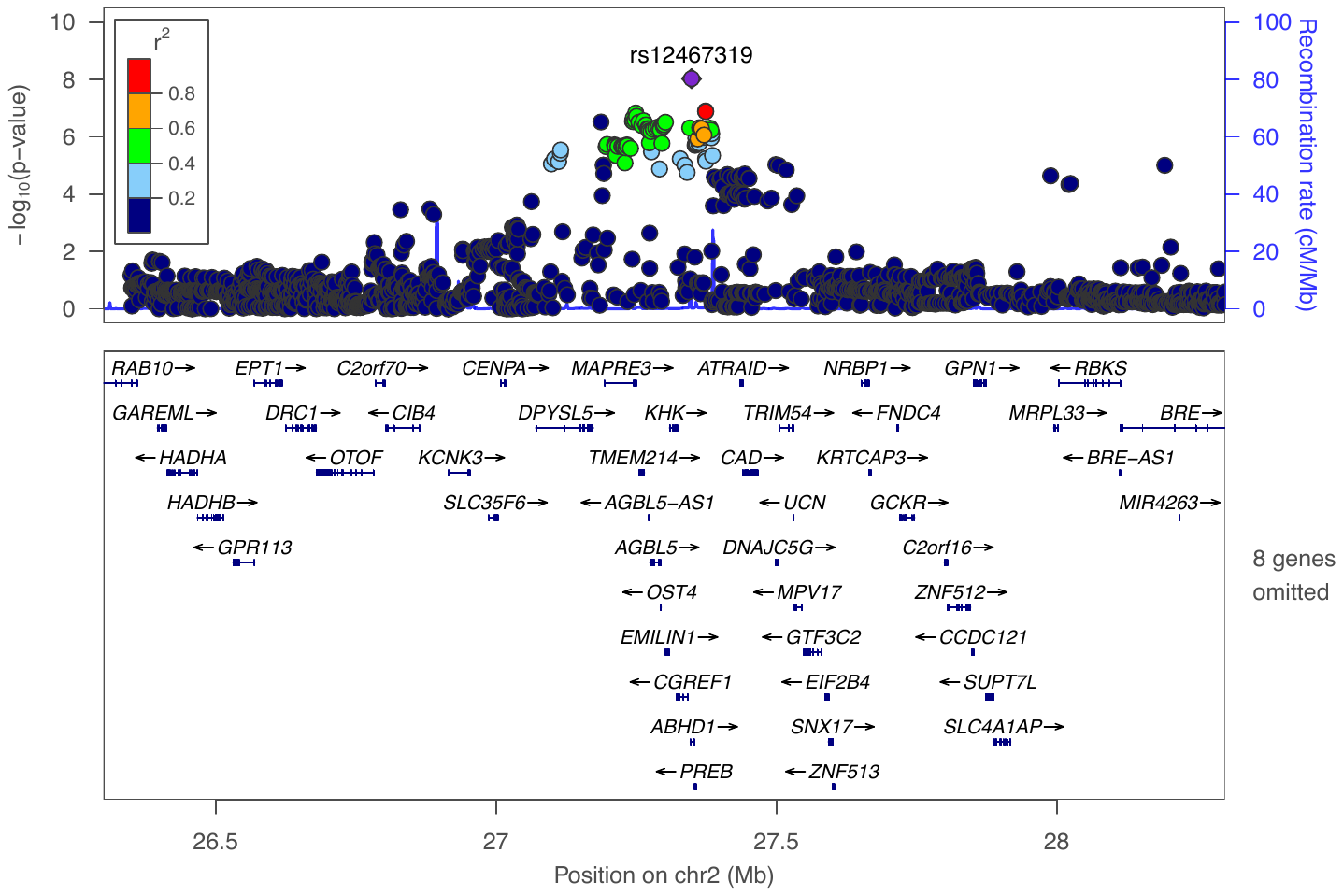

**(b) *TMEM131/*2q11.2**

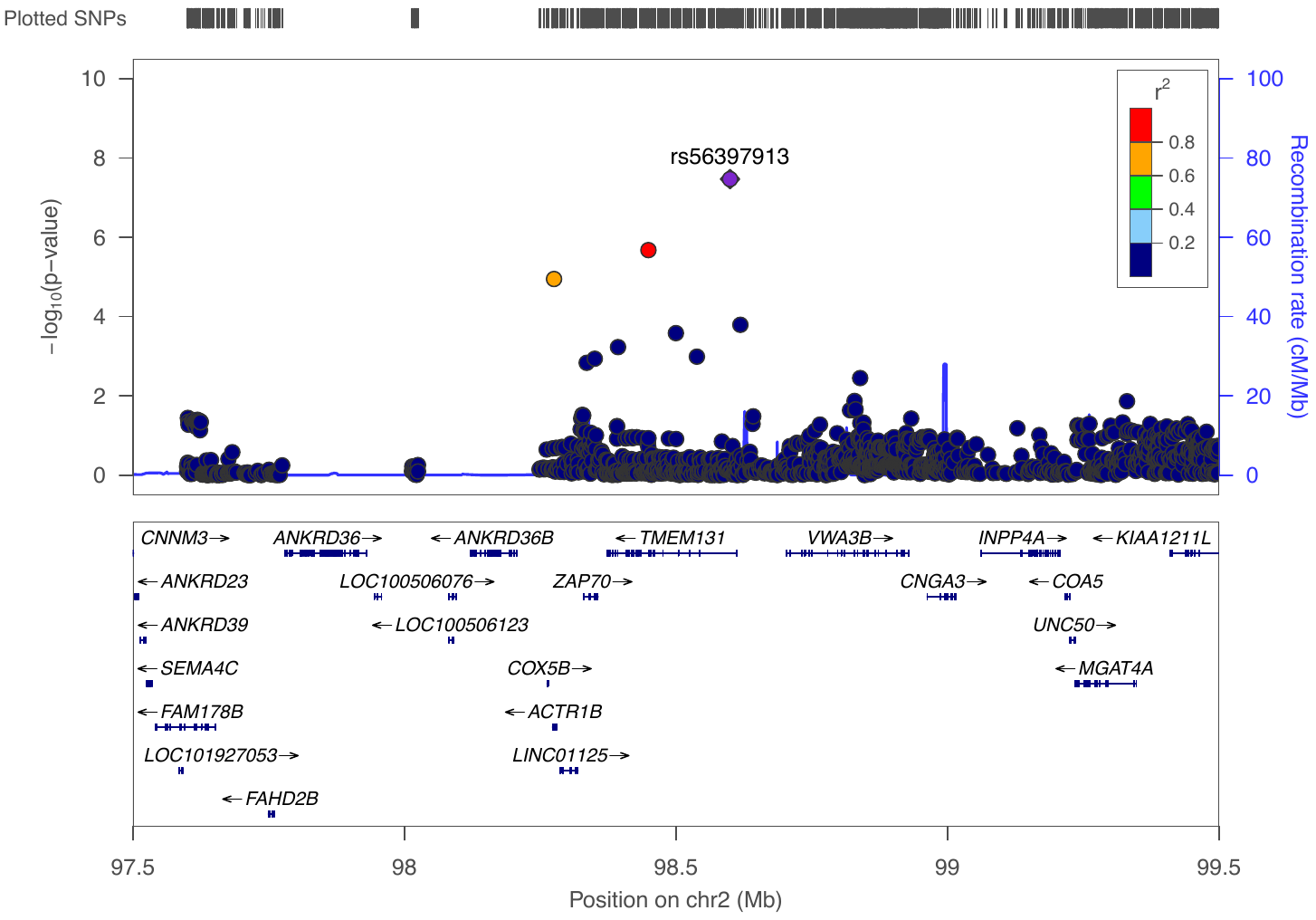

(c) ***XRCC4/*5q14.2**

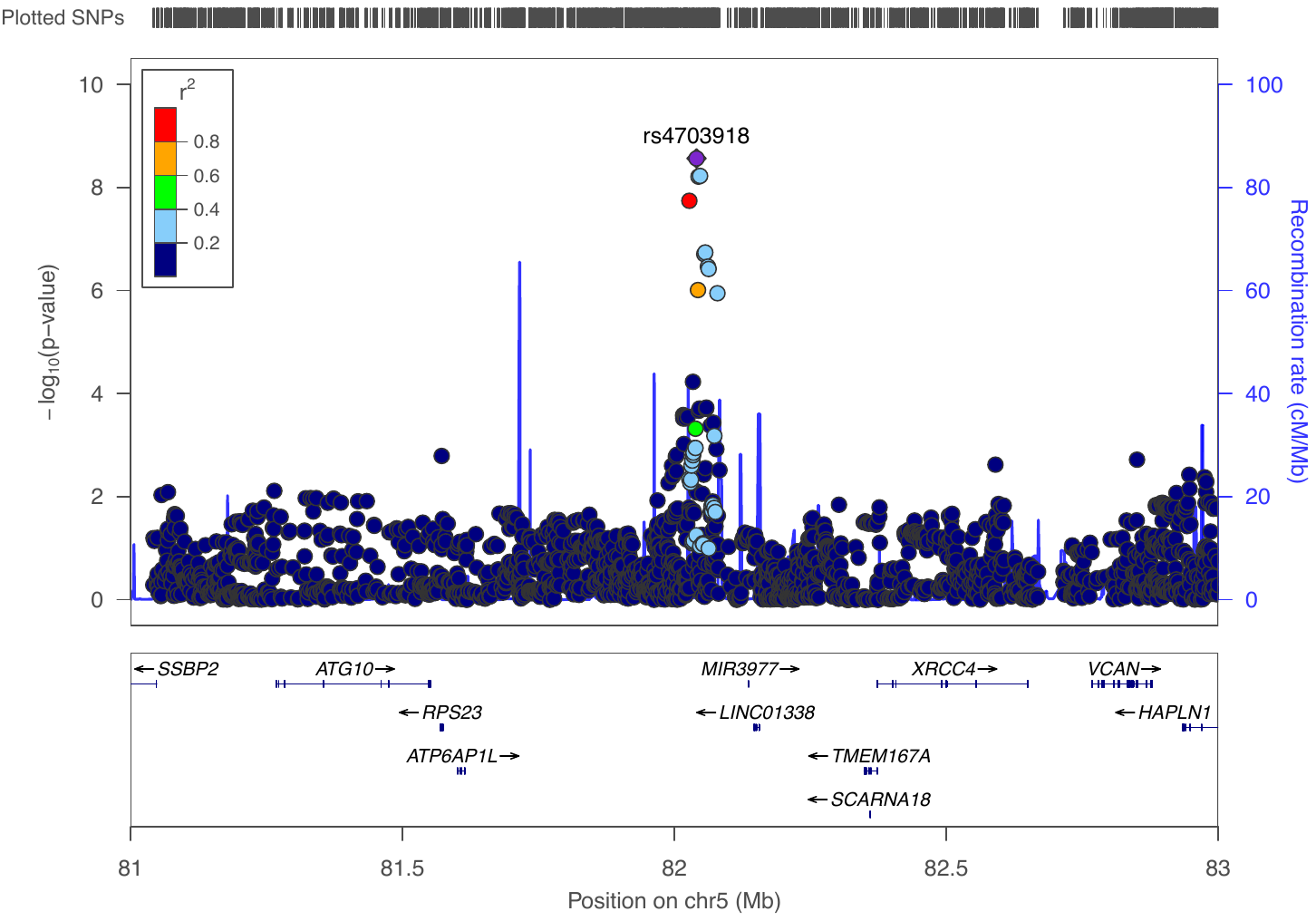

(d) ***PPP1R9A/*7q21.3**

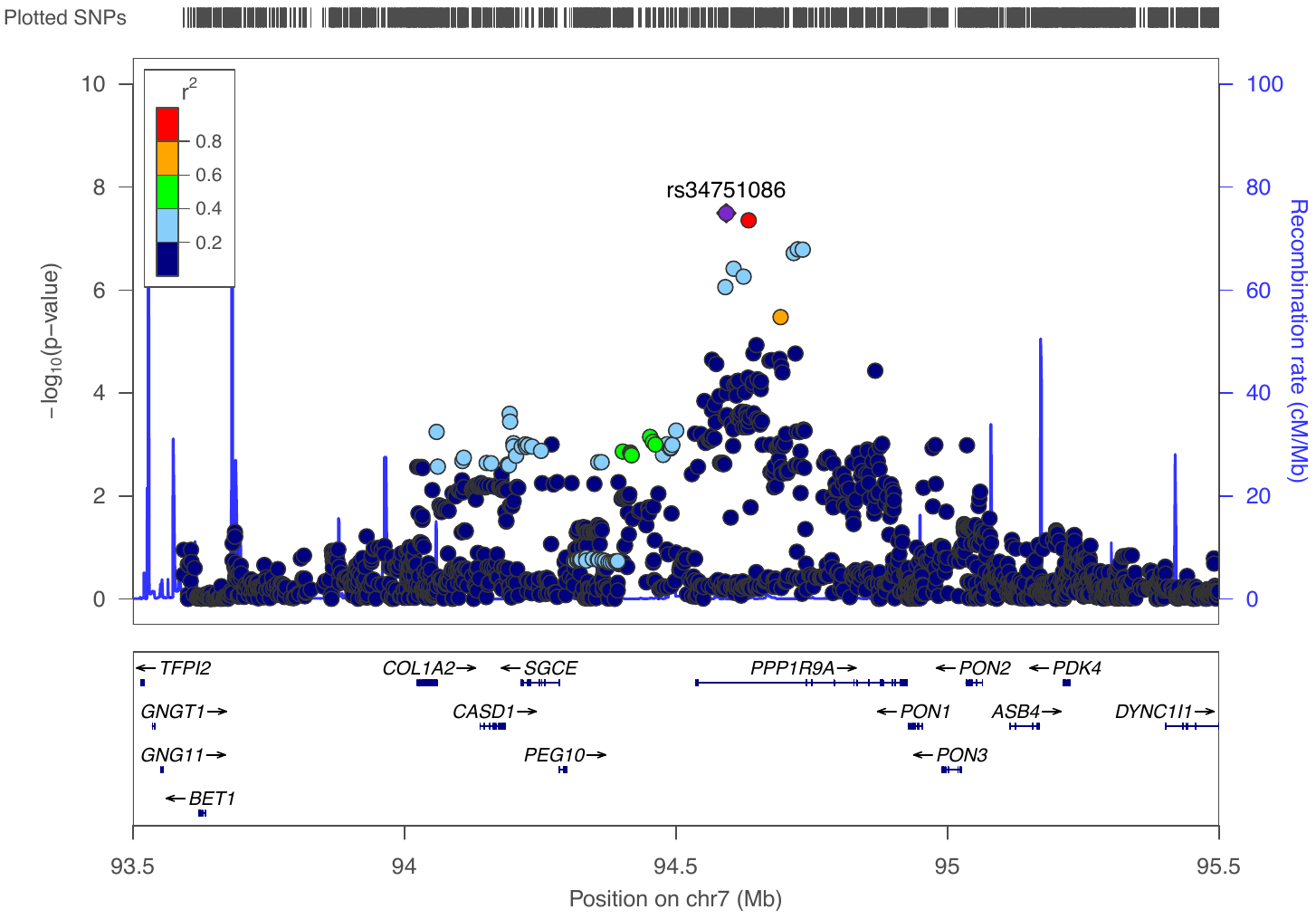

(e) ***XKR6/*8p23.1**

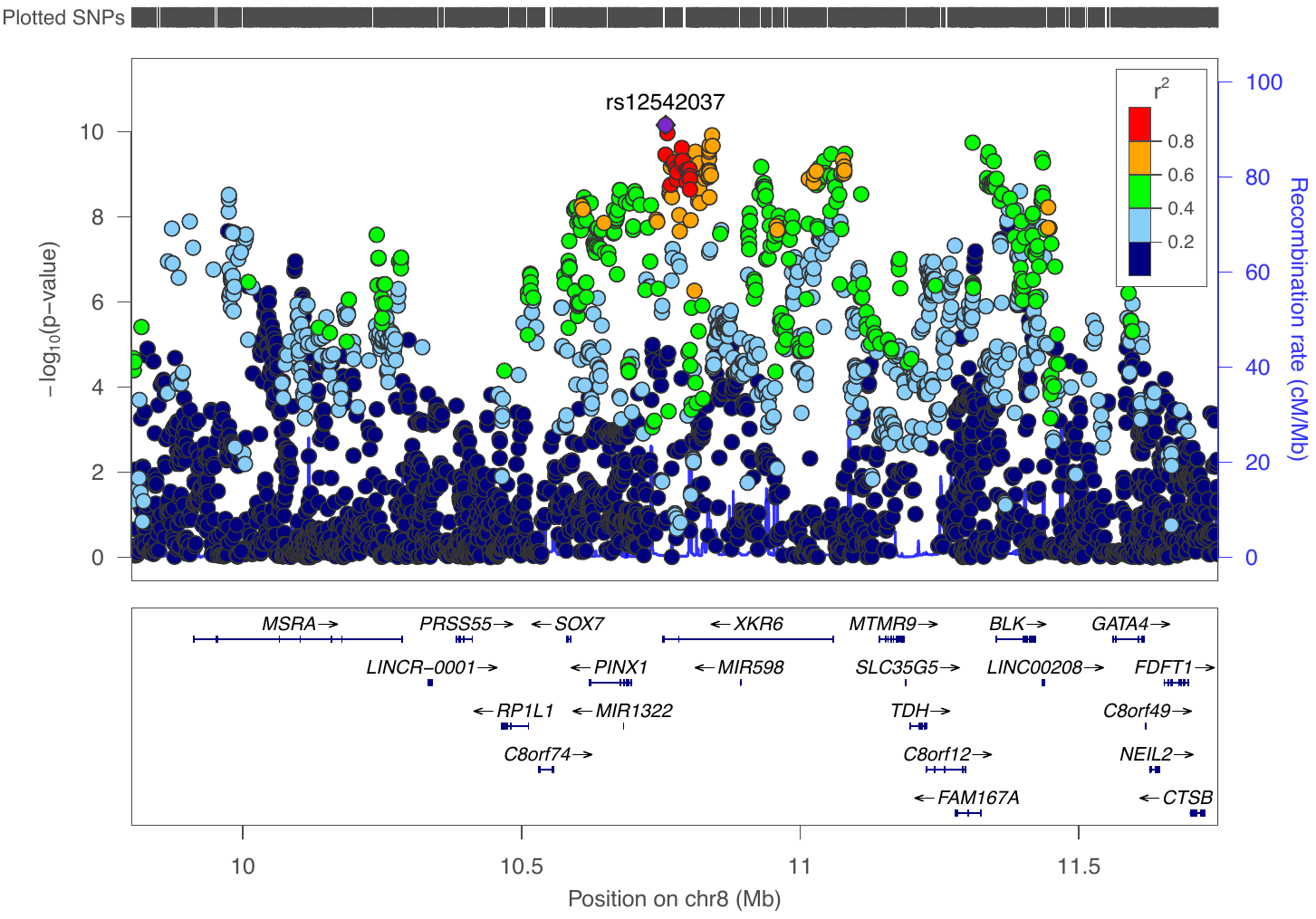

**(f) *TRPS1/*8p23.3**

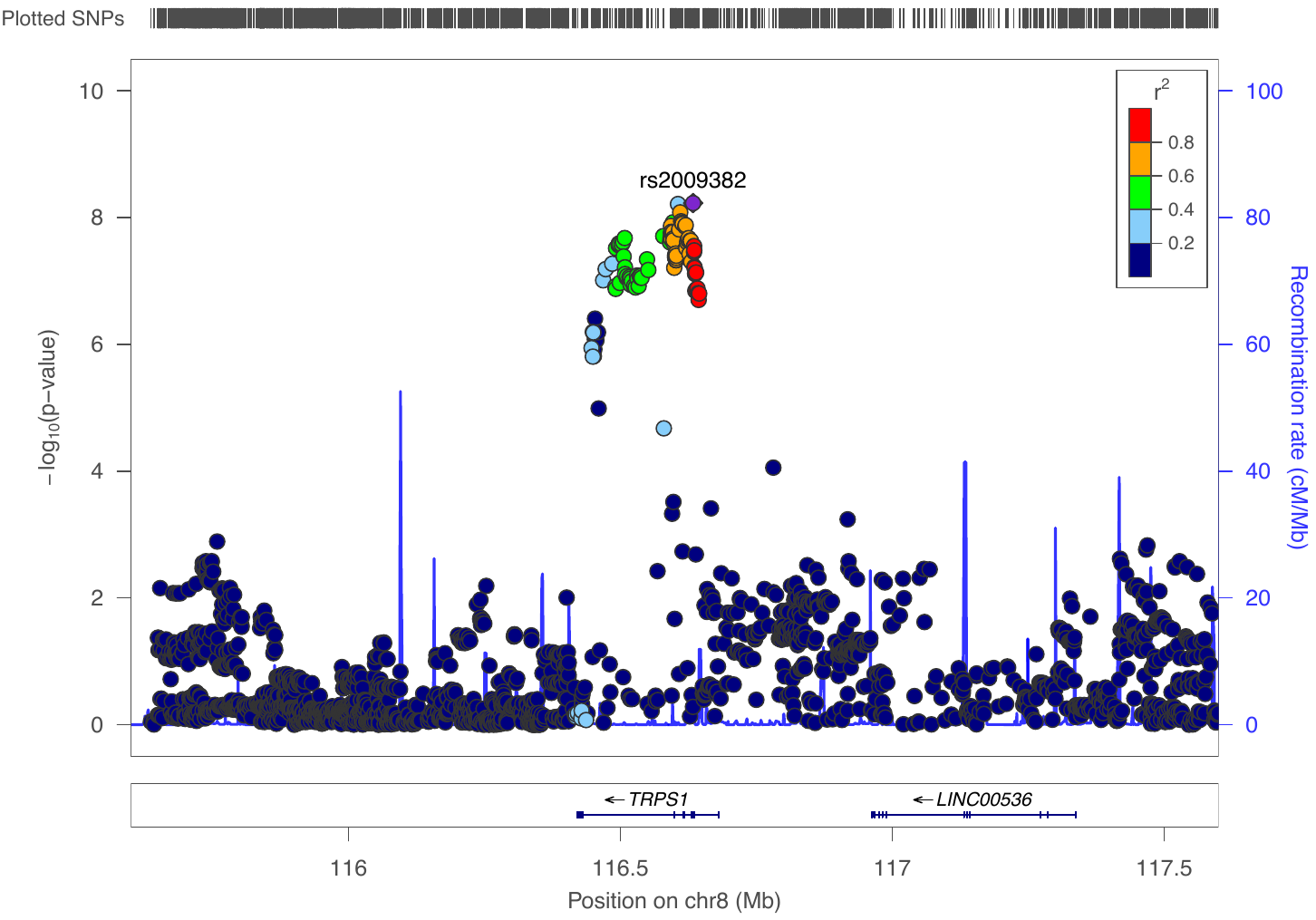

**Supplementary Figure 10.** Top 7 pathways that are enriched with eQTL genes regulated by respective genome-wide significant lead SNPs across the four conditions: endometriosis, osteoarthritis, rheumatoid arthritis and multiple sclerosis.

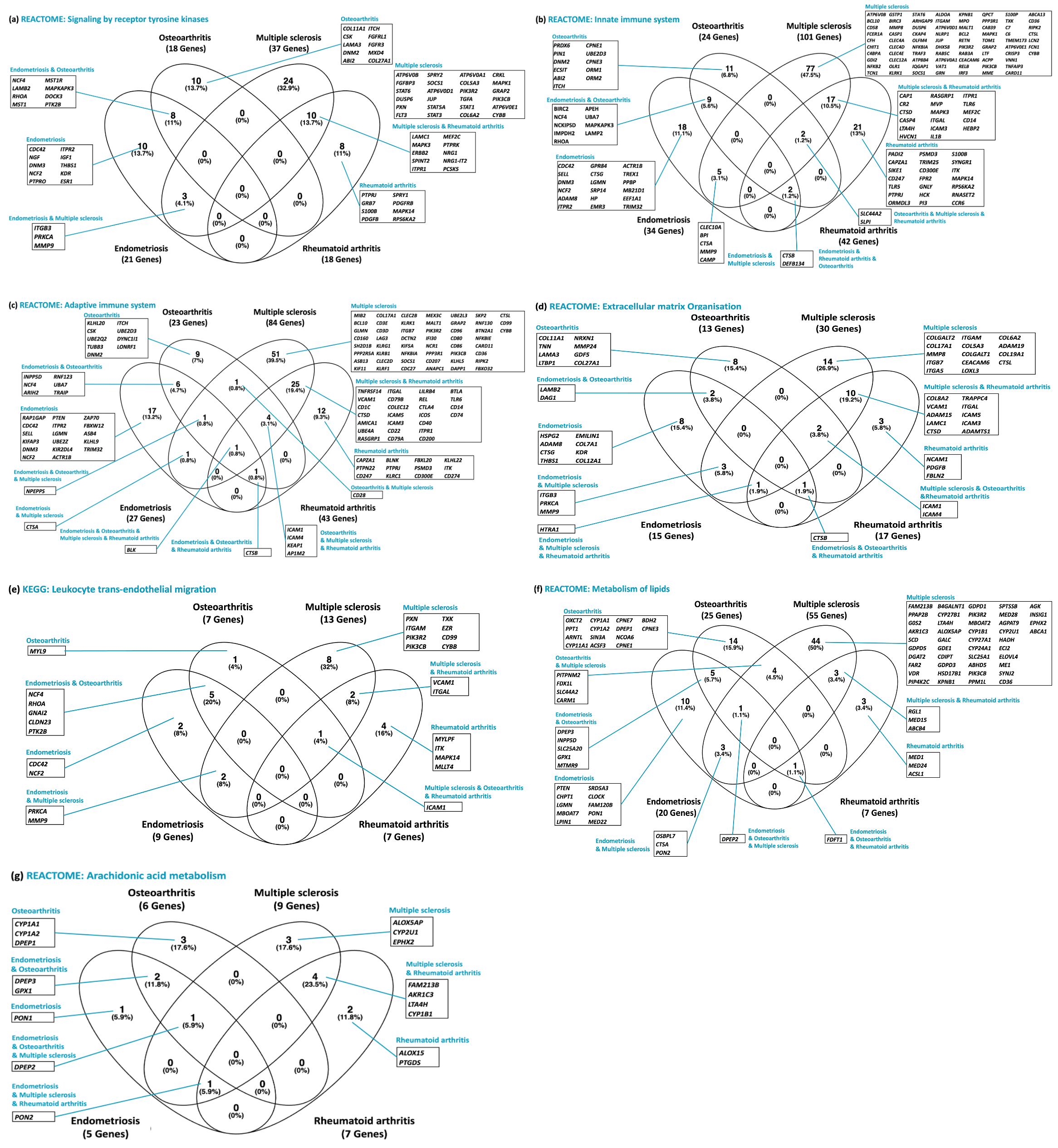
